# International trends in clozapine utilisation prevalence in adults, adolescents and older people: An observational study in 42 countries between 2015–2024

**DOI:** 10.64898/2026.09.17.26363131

**Authors:** Oliver Scholle, Heidi Taipale, Michael Dörks, Jannik Ohmes, Lise Aagaard, Oleg Aizberg, Salim Al Huseini, Mohamed Alnor, Stojan Bajraktarov, Paolo Bertolini, Robert A. Bittner, Syed Ali Bokhari, Alexandra Bražinová, Dagmar Breznoščáková, Kirsten Catthoor, Marko Čavlina, Andreja Čelofiga, Sherry K. W. Chan, Robert O. Cotes, Adomas Danilevičius, Vlad Dionisie, Uwe Eichler, Zsófia Engi, Rifai Farid, Matthäus Fellinger, Kari Furu, Ary Gadelha, Emily L. Griner, Larus S. Guðmundsson, Daniel Guinart, Katarina Gvozdanović, Yuqi Hu, Fuad N. Ismayilov, Jamila Ismayilova, Sung Woo Joo, Luuk J. Kalverdijk, Mulualem Kelebie, Yuki Kikuchi, Nadzeya Kislaya, Miloslav Kopecek, Francisco T. T. Lai, Hsien-Yuan Lane, Jimmy Lee, Jungsun Lee, Soffy C. López, Jorge E. Machado-Alba, Raffael Massuda, Géric Maura, Claude Mawa, Cristián Mena, Georgios Mikellides, Hassan Mirza, Philippe Mortier, Peter Niemegeers, Prasad S. Nishtala, Tīna Nulle, Uladzimir Pikirenia, Gerald J. Pruckner, Maria Puiu, Julieta Ramirez, Johan Reutfors, Ieva Saliete, Fariza Sani, Catharina C.M. Schuiling-Veninga, Dan Siskind, Lina Škiudaitė, Samvel Sukiasyan, Charmaine Tang, Karine Tataryan, David Taylor, Antonio Teixeira Rodrigues, Hiroaki Tomita, Carla Torre, Mike Trott, Fuu-Jen Tsai, Hélène Verdoux, Yahyah Wehbe, Ebenezer Oloyede, Christian J. Bachmann

## Abstract

**Importance:** Clozapine is the only evidence-based treatment for treatment-resistant schizophrenia (TRS), yet substantial international variation in its use has been reported.

**Objective:** To provide a comprehensive and updated assessment of international clozapine utilisation by estimating prevalence across countries using a standardised methodology and examining factors associated with variation in use.

**Design, Setting, and Participants:** Repeated cross-sectional study using population-based data from 2015 to 2024. Individual-level data were obtained from 36 countries and 1 special administrative region, supplemented by national clozapine consumption data from 5 additional countries. Analyses were conducted separately for adults (20-64 years; primary analysis), adolescents (10-19 years), and older adults (≥65 years).

**Main Outcomes and Measures:** Annual outpatient clozapine utilisation prevalence per 100,000 population by age group, sex, country and calendar year. Associations between clozapine utilisation prevalence and country-level health system, demographic, and socioeconomic indicators were also examined.

**Results:** We analysed data from more than 1.1 billion people across all inhabited continents. In 2024, crude adult clozapine use prevalence was highest in Croatia (415.3 per 100,000 population) and Finland (302.0 per 100,000), and lowest in Oman (0.5 per 100,000) and Ethiopia (0.3 per 100,000), representing a 1,384-fold difference. Across most countries, utilisation peaked among individuals aged 50 to 59 years. The male-to-female prevalence ratio ranged from 0.4 in the United Arab Emirates to 2.8 in Colombia. Among adolescents, prevalence was highest in Colombia (42.7 per 100,000) and Croatia (24.4 per 100,000), while among older adults it was highest in Croatia (406.9 per 100,000) and Iceland (339.7 per 100,000). Across all countries, adult clozapine utilisation increased by 17% (95% CI, 16%-18%), from 44.7 per 100,000 population (95% CI, 44.5-44.9) to 52.2 per 100,000 population (95% CI, 51.1-52.4). Country-specific trends ranged from a 71% decrease to a 342% increase. None of the examined indicators explained variation in clozapine utilisation across countries.

**Conclusion and relevance:** Clozapine use increased modestly over the past decade but remained highly variable between countries. This variation was not explained by national socioeconomic, healthcare-system, or monitoring-related factors, suggesting an important role for clinical and service-level barriers. The concentration of clozapine use in later adulthood is consistent with delayed initiation and highlights opportunities to improve timely access to evidence-based treatment for TRS.

**Funding:** Hsien-Yuan Lane: National Science and Technology Council, Taiwan (NSTC 114-2629-B-039-001-); The China Medical University Hospital Research Project (DMR-113-009 and DMR-114-046). Philippe Mortier: Miguel Servet grant CP21/00078 co–financed by the Instituto de Salud Carlos III (ISCIII) and co–funded by the European Union; grant PI22/00107 funded by ISCIII and co–funded by the European Union; and grant 202220–30–31 from the Fundació la Marató de TV3. Heidi Taipale: Sigrid Jusélius Foundation grant.

## Introduction

Clozapine is the only antipsychotic recommended for the treatment of treatment-resistant schizophrenia (TRS), broadly defined as an inadequate response to at least two adequate trials of antipsychotic medication.^1^ TRS affects approximately one-third of individuals diagnosed with schizophrenia and is associated with substantial morbidity and premature mortality.^2,3^ Reflecting its superior effectiveness, clozapine constitutes part of the WHO Model List of Essential Medicines.^4^

Following its initial clinical introduction in the early 1970s, clozapine was withdrawn in the United States and most European countries after reports of fatal agranulocytosis, most notably eight deaths in Finland.^5^ Despite these early safety concerns, clozapine was subsequently reintroduced due to its superior clinical efficacy, accompanied in most regions by mandatory haematological monitoring requirements.^6,7^ These regulatory responses, shaped largely by the Finnish experience, have had a large influence on clinical practice and perceptions of clozapine treatment internationally.

Substantive evidence demonstrates that clozapine is more effective than other antipsychotics for reducing psychotic symptoms,^8,9^ suicidality,^10,11^ violent behaviour,^12^ and all-cause mortality in schizophrenia.^13,14^ In addition to its established efficacy in schizophrenia, emerging evidence suggests that clozapine may confer broader transdiagnostic benefits,^15^ including reduced psychiatric hospitalisations^16,17^ and favourable health economic effects.^18,19^ Nevertheless, clozapine remains markedly underutilised among patients who are likely to benefit.^20^ Delayed initiation and non-prescription are thought to be multifactorial, with commonly cited barriers including the burden of routine blood monitoring, prescriber inexperience, concerns regarding adverse effects, institutional constraints and assumptions about patient unwillingness to engage with monitoring requirements.^21^

Although clozapine-induced agranulocytosis is a potentially fatal adverse event, its risk is time-limited, with incidence peaking early in treatment and declining substantially thereafter from approximately 0.13% at nine weeks to around 0.001% after two years^22^, or from an adjusted odds ratio of 36 within the first six months to 4 after four to five years of treatment.^23^ Importantly, patients frequently report high treatment satisfaction with clozapine despite ongoing adverse effects.^24^ Moreover, several other serious clozapine-associated complications, including gastrointestinal hypomotility, myocarditis, cardiomyopathy, seizures and hypersalivation, carry mortality risks comparable to or greater than that of agranulocytosis, underscoring the importance of balanced risk appraisal.^25,26^

International variation in clozapine utilisation prevalence has been described previously.^20,27,28^ However, as prescribing practices, clinician experience, service organisation and attitudes towards clozapine evolve over time, it remains unclear how utilisation prevalence has changed across countries and regions in recent years. The aim of this study was therefore to provide an updated and comprehensive overview of international clozapine utilisation prevalence and temporal trends, using a consistent analytical protocol, with a primary focus on adults aged 20–64 years, the population most relevant to clozapine treatment for TRS. Generating comparable international estimates is essential for identifying gaps in care and informing targeted, system level interventions to improve access to this highly effective treatment.

## Methods

We conducted a cross-national comparison drug utilisation study to compare clozapine use across multiple countries.

### Study Design

Using a common protocol, we applied a repeated cross-sectional design to population-based data from 41 countries and one Special Administrative Region (SAR). The participating countries/SAR were Argentina, Armenia, Australia, Austria, Azerbaijan, Belarus, Belgium, Brazil, Brunei, Chile, Colombia, Croatia, Cyprus, Czech Republic, Denmark, Ethiopia, Finland, France, Germany, Hong Kong SAR, Hungary, Iceland, Japan, Latvia, Lithuania, Netherlands, New Zealand, North Macedonia, Norway, Oman, Poland, Portugal, Romania, Singapore, Slovakia, Slovenia, South Korea, Spain, Sweden, Taiwan, the United Arab Emirates, and the United States of America. Details on the data sources for each country/SAR can be found in **Table S1**.

### Data collection

Participating countries/SAR were requested to provide population-based data for the calendar years 2015–2024 (where available). All requested information was specified a priori to enable harmonised analyses; the primary authors received only aggregated data, while access to individual-level data remained restricted to country-level investigators, or clozapine register/data source holders. The study population was intended to be as representative of the general population as possible and comprised individuals of all ages; analyses were restricted to female and male sex, as other gender identities could not be included due to their expected low frequency and limited data availability across sources.

For each country/SAR and calendar year, stratified by sex and 5-year age groups (0–4, 5–9, …, 75–79, 80+ years), the requested data comprised the absolute number of individuals in the study population and the absolute number who received at least one prescription and/or dispensation of clozapine. Clozapine was identified using the WHO Anatomical Therapeutic Chemical (ATC) code N05AH02.^29^ For some countries where the requested standard data set was not fully available, deviations from the standard data set were accepted. These included the use of alternative age bands, no stratification by sex and age, or reporting defined daily doses (DDDs; current DDD for clozapine: 300 milligrams per day)^29^ in place of prescription or dispensing prevalence. Data provided under these specifications were reported to maximise coverage and cross-national comparisons.

### Data analysis

We calculated the annual clozapine utilisation prevalence per 100,000 population by dividing the number of individuals who received at least one prescription and/or dispensation of clozapine by the total number of individuals in the study population for the corresponding calendar year, multiplied by 100,000. Clozapine utilisation prevalence was calculated for adults (main analysis; 20–64 years), adolescents (10–19 years) and older adults (≥65 years). Because we primarily report crude prevalence, we additionally age- and sex-standardised overall clozapine utilisation prevalence for countries with available age- and sex-stratified data, using the European Standard Population (Eurostat, 2013 revision)^30^ as a reference. The standardised estimates were used to assess the impact of standardisation and in analyses of relative changes between the earliest and the latest calendar year of the study period.

We further conducted exploratory cross-national analyses of the association between crude clozapine utilisation prevalence for the age group 20–64 years in 2024 (or the most recent year available) and a panel of country-level health system and socioeconomic indicators. These analyses were intended to descriptively explore potential patterns across countries and to generate hypotheses rather than to support causal inference. Associations between each indicator and clozapine utilisation prevalence were assessed separately using Pearson’s correlation coefficient, assuming an approximately linear relationship between variables. Statistical uncertainty was described using 95% confidence intervals and two-sided p-values based on the t-distribution with n–2 degrees of freedom, where n denotes the number of paired observations. Given the ecological and exploratory nature of these analyses and the limited number of countries, no multivariable modelling was performed. The indicators included population size (data source: United Nations, World Population Prospects), degree of urbanisation (World Factbook Urbanisation), GDP per capita (World Bank), GINI coefficient (World Bank Poverty and Inequality Platform), health expenditure (percentage of GDP; World Bank), health expenditure (per capita; World Bank), mental health spending (percentage of health budget; WHO), psychiatrist density (number of psychiatrists per 100,000 inhabitants; WHO) and the clozapine monitoring stringency index.^7^ The statistical analyses were performed with Excel 2024 (Microsoft Corporation; Redmond, WA, USA).

### Ethical approval

Where applicable, the contributing authors sought ethics committee or institutional review board approval (**Table S2**).

## Results

Overall, 42 countries/SAR provided data and were included in the study (**Table S1**), with 37 countries/SAR providing aggregated individual-level data on a total of 1,181,534,866 persons (most recent data year). Of these, 31 supplied clozapine utilisation prevalence data stratified by age and sex, which formed the basis for the main analysis of adults aged 20–64 years. Six countries reported prevalence data for the total population only. An additional five countries reported solely total DDD per population. A small number of countries deviated from the study period of 2015 to 2024. Data began in 2016 for the USA and 2018 for the United Arab Emirates. The latest available year was 2023 for Austria, South Korea, and the USA and 2022 for Taiwan.

### Clozapine use in adults and variation between countries

In 2024, crude clozapine utilisation prevalence in adults aged 20–64 years was highest in Croatia (415.3/100,000) and in Finland (302.0/100,000), and lowest in Oman (0.5/100,000) and in Ethiopia (0.3/100,000), reflecting up to a 1,384-fold difference between highest and lowest using countries (**Figure 1, Table S3**).

**Figure 1:**
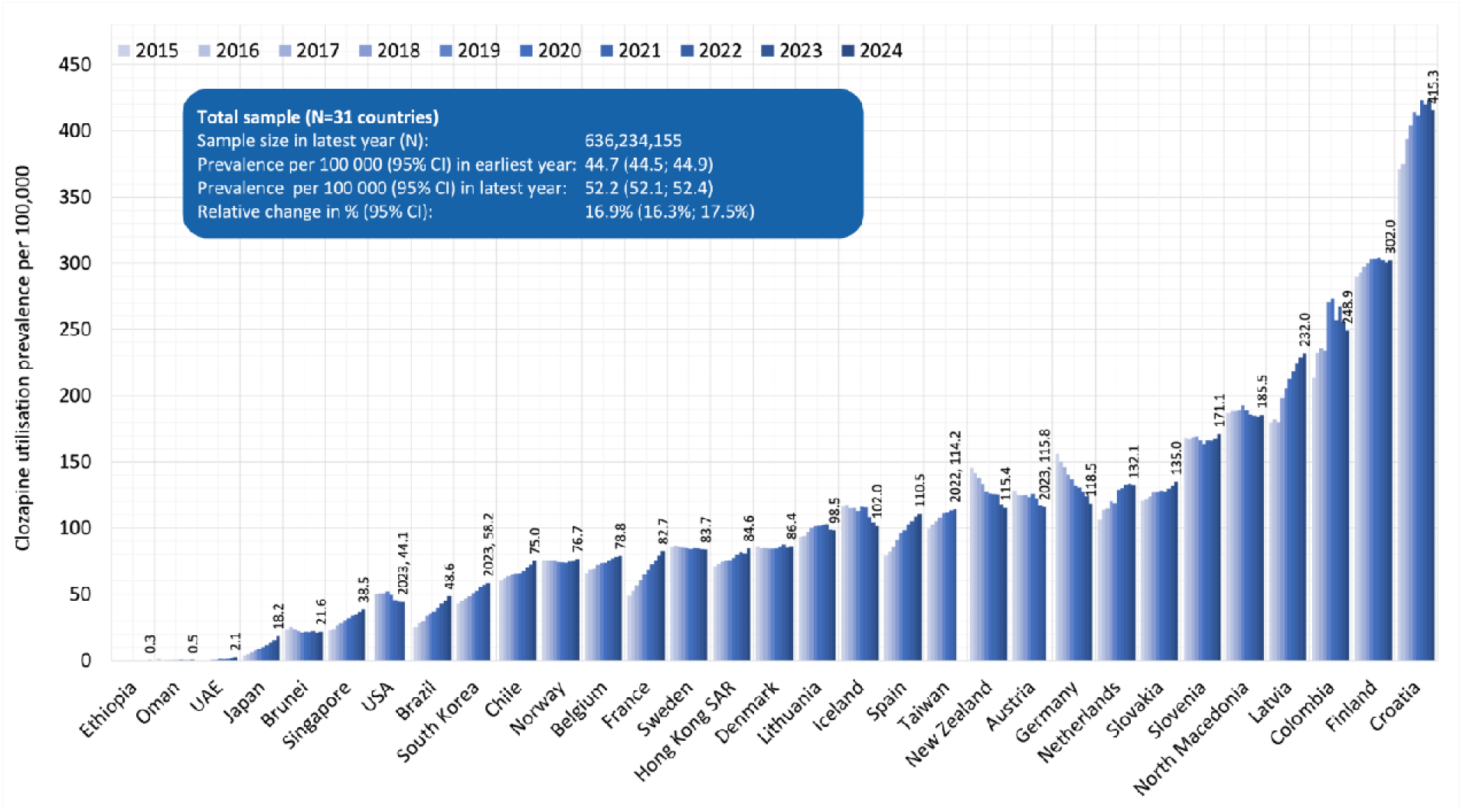
Clozapine utilisation prevalence per 100,000 adults (20–64 years), 2015–2024. SAR = special administrative region; UAE = United Arab Emirates; USA = United States of America.

When using the age- and sex-standardised prevalence, the range of overall clozapine utilisation prevalence narrowed slightly, spanning from 0.3/100,000 in Ethiopia to 406.3/100,000 in Croatia (maximum difference: 1,354-fold) (**Table 1, Figure S1**). The ranking of countries by utilisation prevalence remained largely unchanged.

**Table 1:**
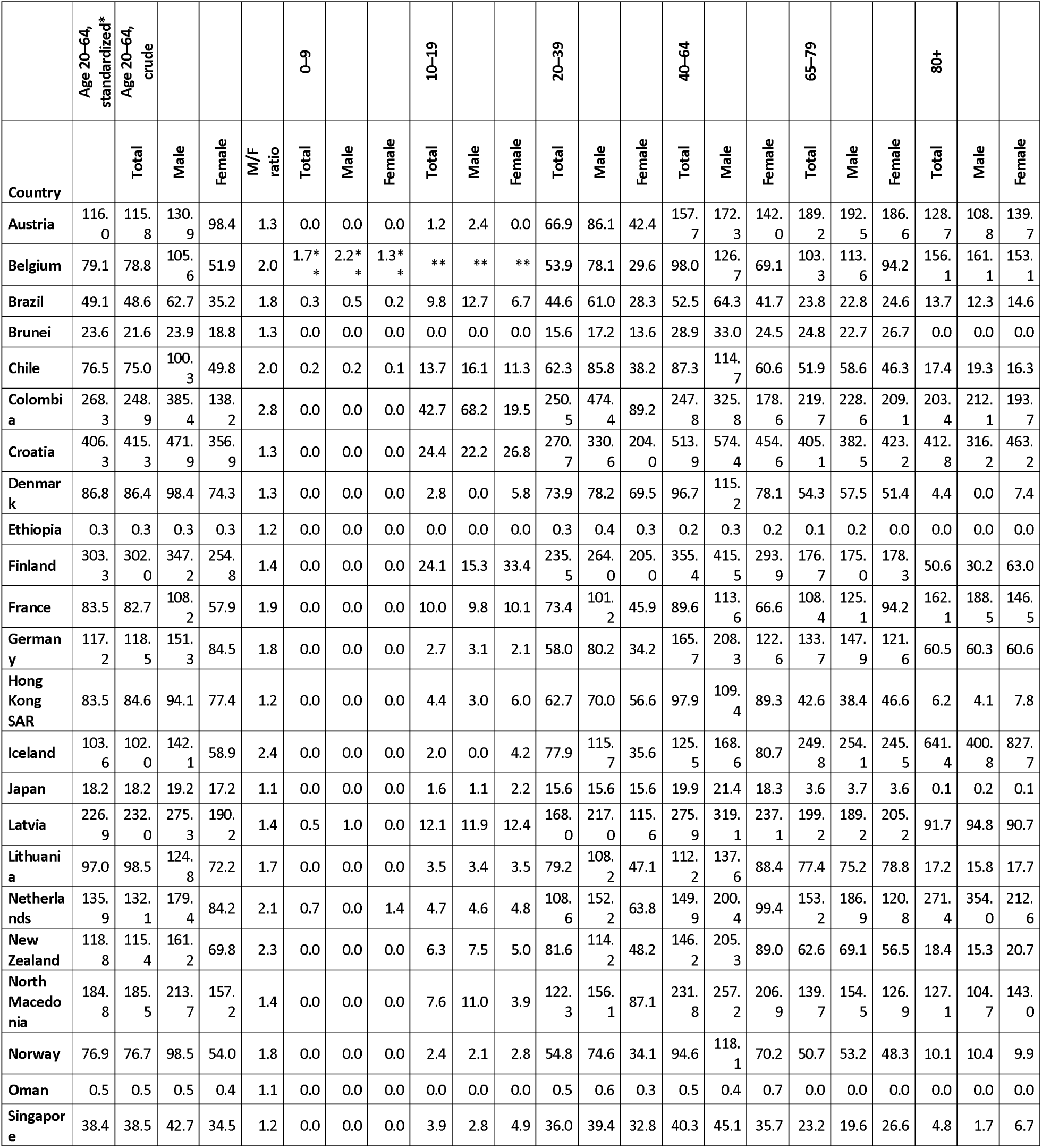
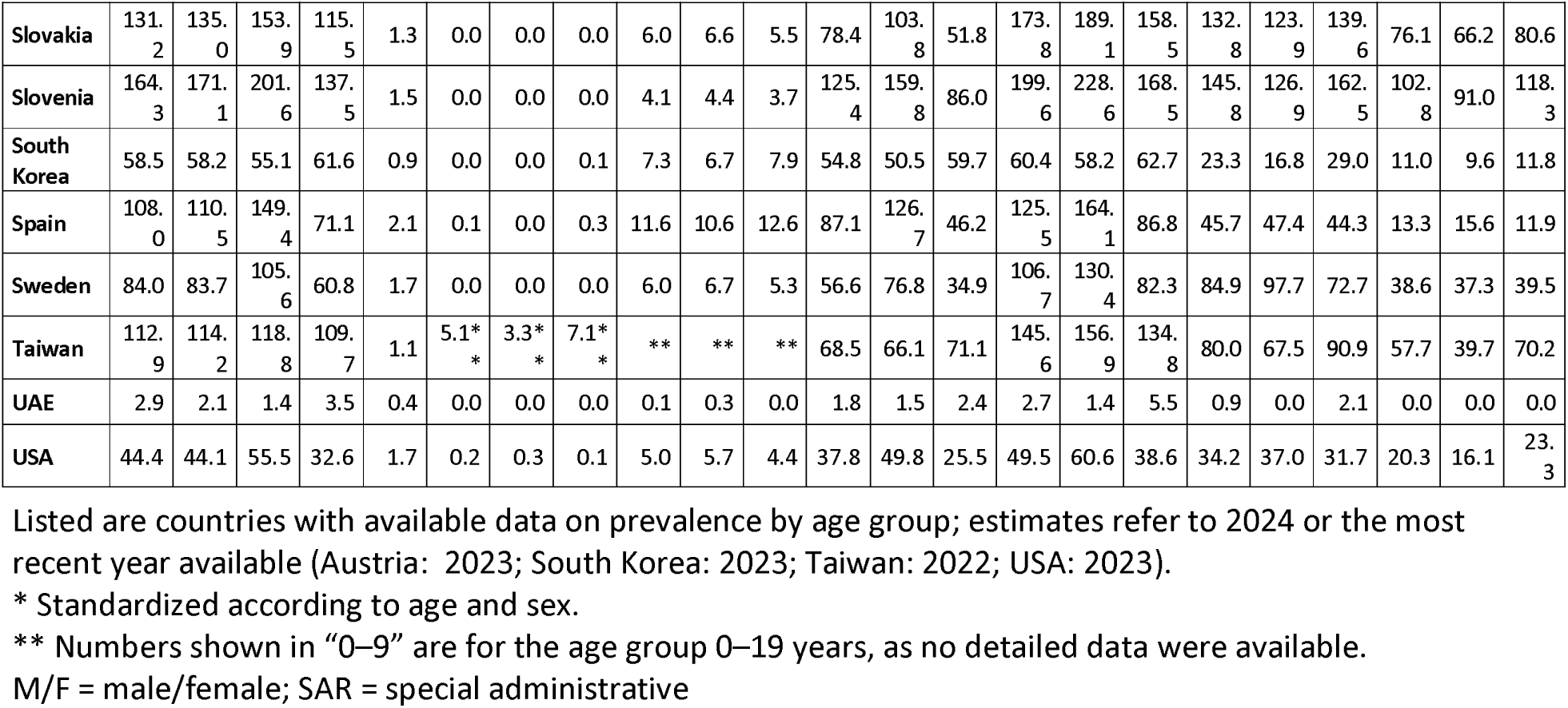
Clozapine utilisation prevalence per 100,000 persons in 2024 (or most recent year available) per country, by sex and age.

### Time trends in clozapine use

Over time, the age- and sex-standardized clozapine utilisation prevalence increased in 16 out of the 31 countries (52%), remained stable (i.e. relative change between −10% and +10%) in nine countries (29%) and decreased in six countries (19%) (**Figure 2**). Relative change in clozapine utilisation prevalence between 2015 and 2024 ranged from increases of +341.7% in Japan and +277.8% in the United Arab Emirates to decreases of −24.5% in Germany, and −71.2% in Oman. The greatest absolute growth in (age- and sex-standardized) clozapine utilisation prevalence over the studied period were observed in Latvia (+44.8/100,000) and Croatia (+37.5/100,000). The largest absolute decrease occurred in New Zealand (−30.7/100,000), and in Germany (−38.1/100,000).

**Figure 2:**
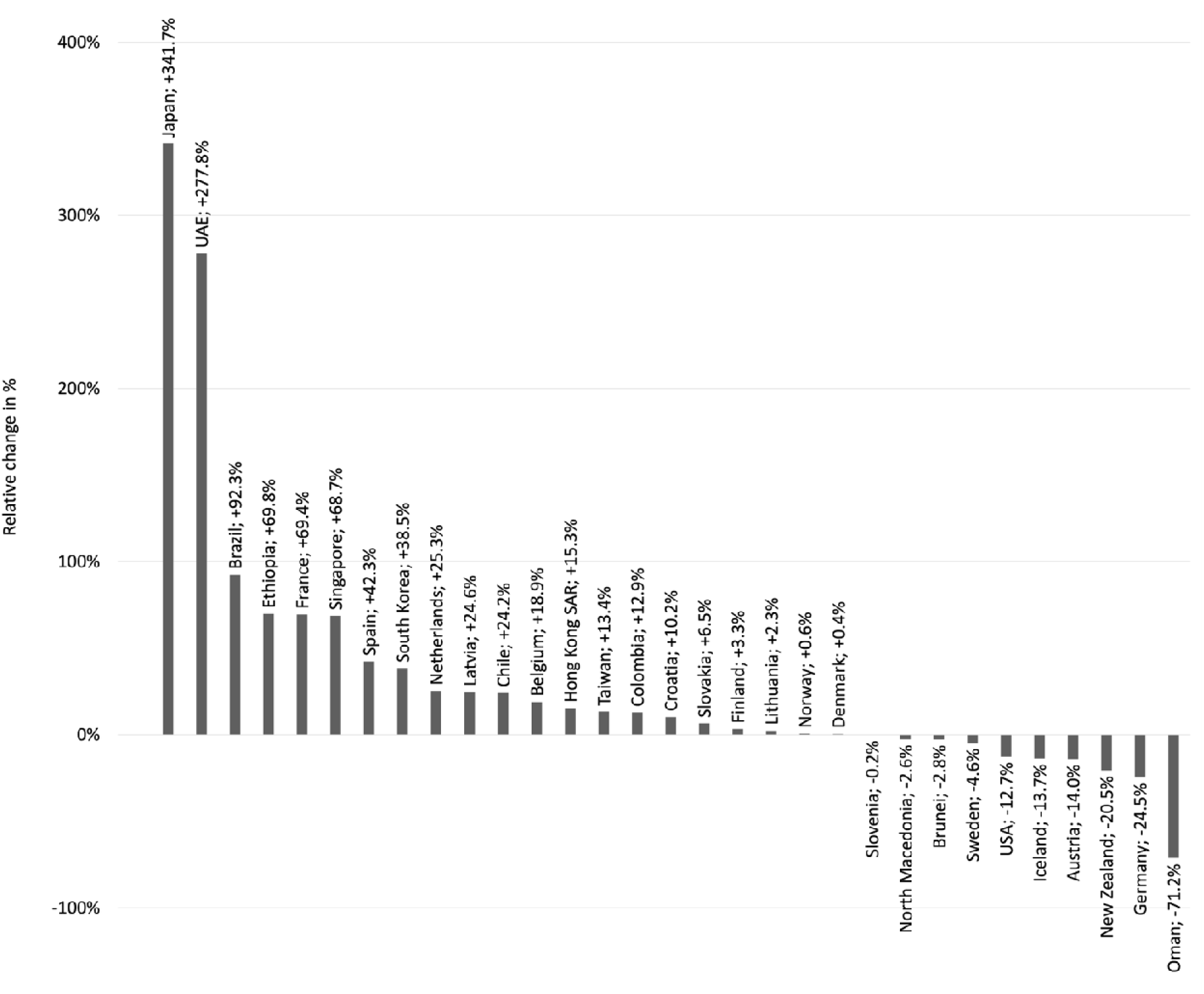
Relative change in age- and sex-standardized clozapine utilisation prevalence in adults (20–64 years) over the study period. SAR = special administrative region; UAE = United Arab Emirates; USA = United States of America.

### Clozapine use by age and sex

**Figure S2** shows clozapine utilisation prevalence in 2024 stratified by age and sex for each country. Across most countries, clozapine utilisation among adults peaked in individuals aged 50 to 59 years. The highest clozapine utilisation prevalence among adults was observed in Croatia (females: 516.1/100,000 in 60- to 64-year-olds; males: 621.7/100,000 in 50- to 54-year-olds). In most countries (29 of 31; 94%), clozapine use was higher in males than in females, with the male/female ratio ranging from 0.4 (United Arab Emirates) to 2.8 (Colombia) (**Table 1**).

### Association between clozapine utilisation and country-level socio-medico-economic indicators

Regarding potential influencing factors, no associations were observed between clozapine utilisation prevalence in adults and countries’ gross domestic product, GINI coefficient, degree of urbanisation, population size, general health expenditures, mental health spending, psychiatrist density, or clozapine monitoring stringency (**Table 2**).

**Table 2:**
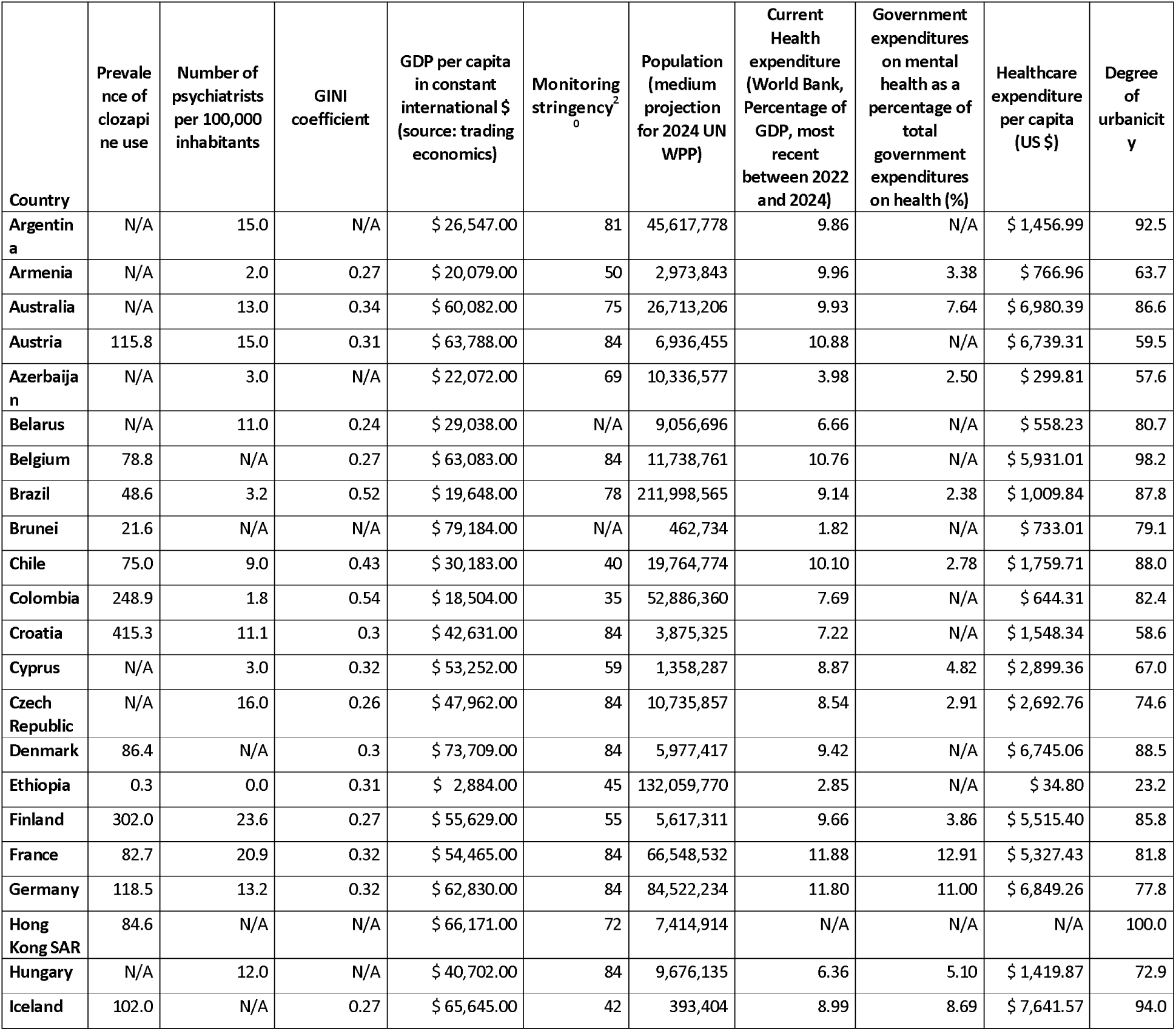
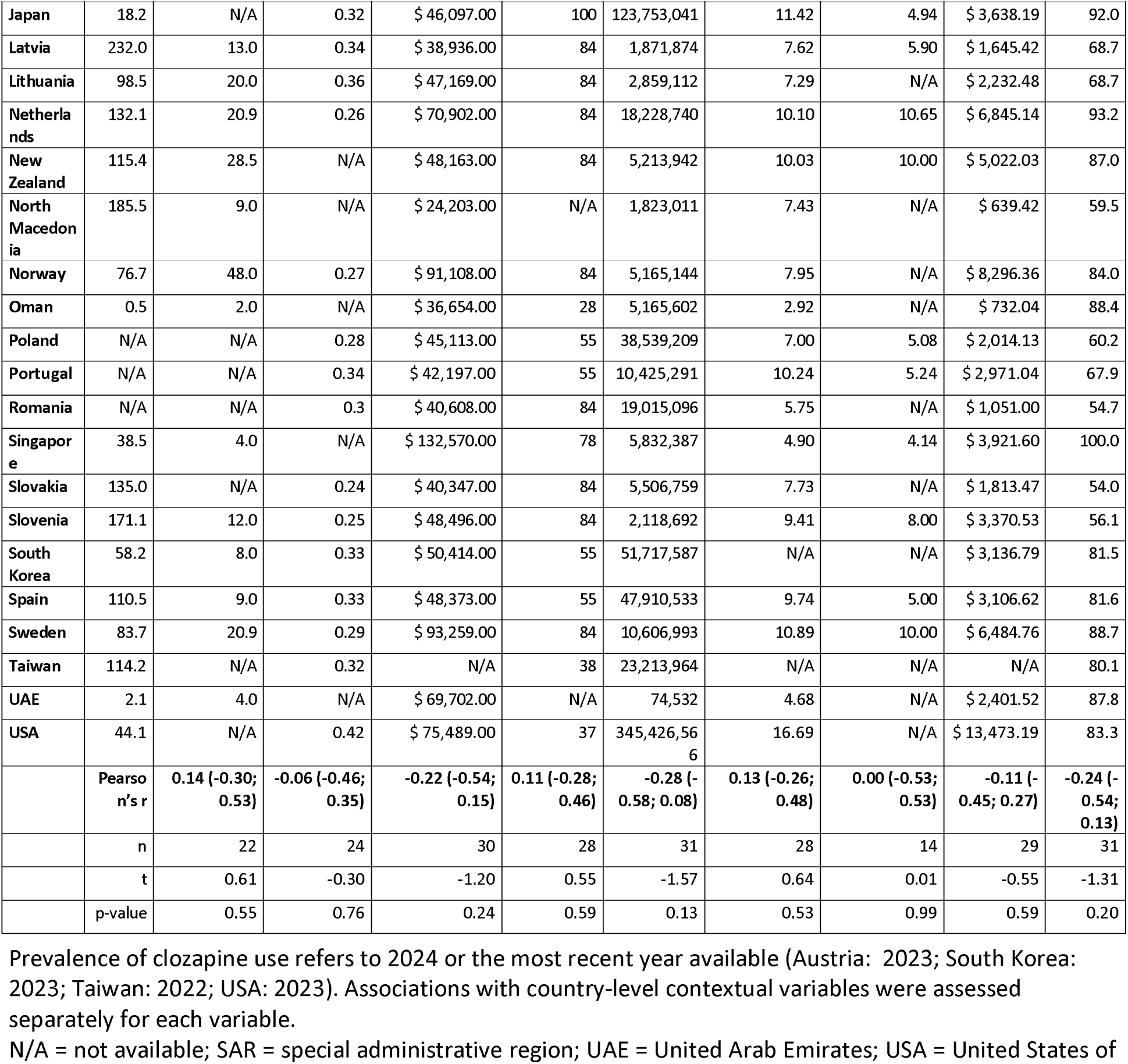
Univariate associations between clozapine utilisation prevalence and country-level indicators.

### Clozapine use in adolescents and in older adults

In 2024, clozapine use among adolescents (10–19 years) was highest in Colombia (42.7/100,000) and in Croatia (24.4/100,000) (**Figure 3**). Across the study period, clozapine utilisation prevalence in adolescents increased (i.e., relative change ≥+10%) in 12 out of the 29 countries (41%) (**Figure 3; Table S4**).

**Figure 3:**
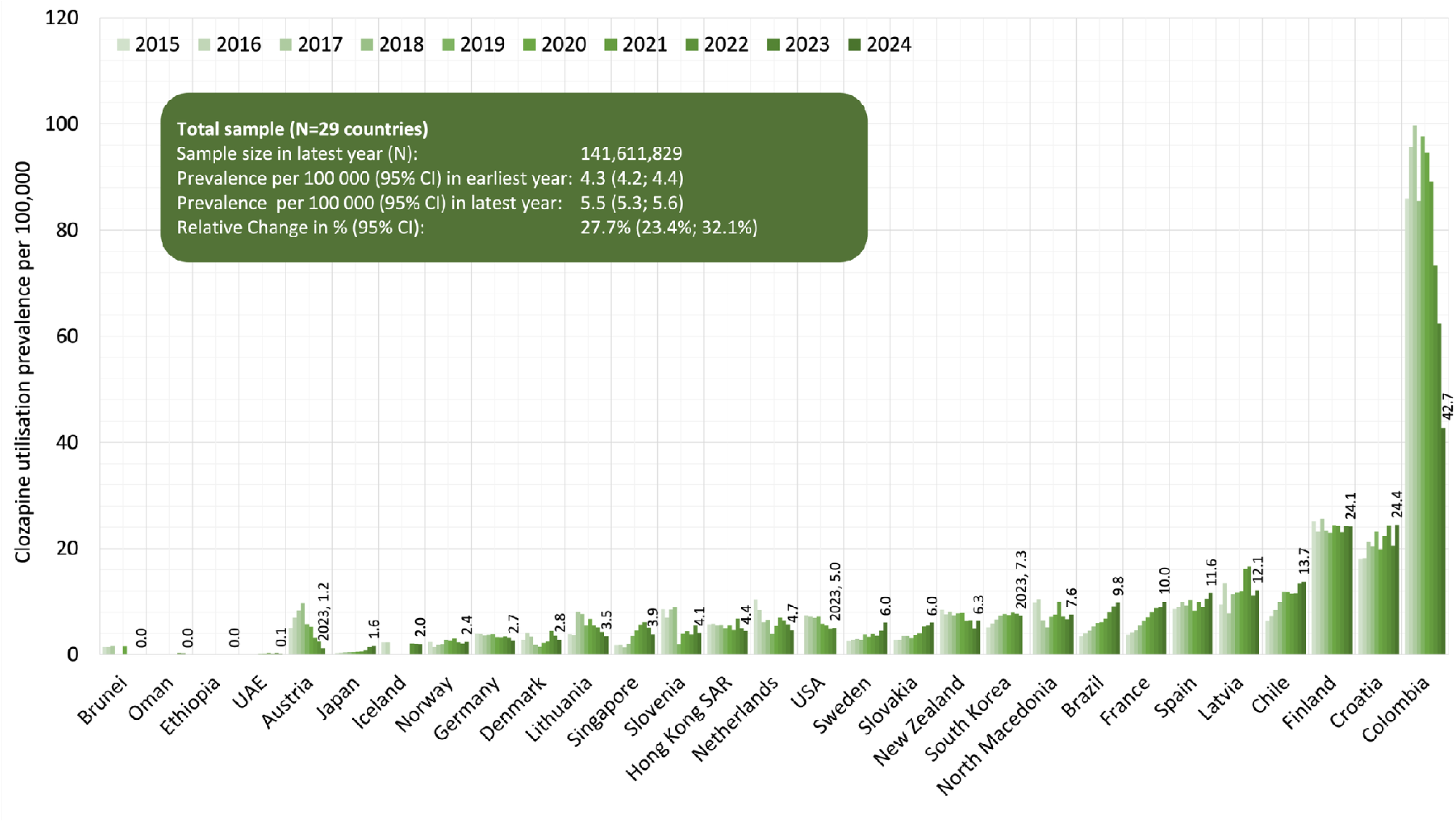
Clozapine utilisation prevalence per 100,000 adolescents (10–19 years), 2015–2024. SAR = special administrative region; UAE = United Arab Emirates; USA = United States of America.

Among older adults (≥65 years), clozapine utilisation prevalence in 2024 was highest in Croatia (406.9/100,000) and Iceland (339.7/100,000) (**Figure 4; Table S5**), with a particularly high prevalence of clozapine utilisation among Icelandic women aged 80 years and older, exceeding 800 per 100,000 (**Table 1**). Across the study period, clozapine utilisation in this age group increased in 18 of the 31 countries (58%) (**Figure 4**).

**Figure 4:**
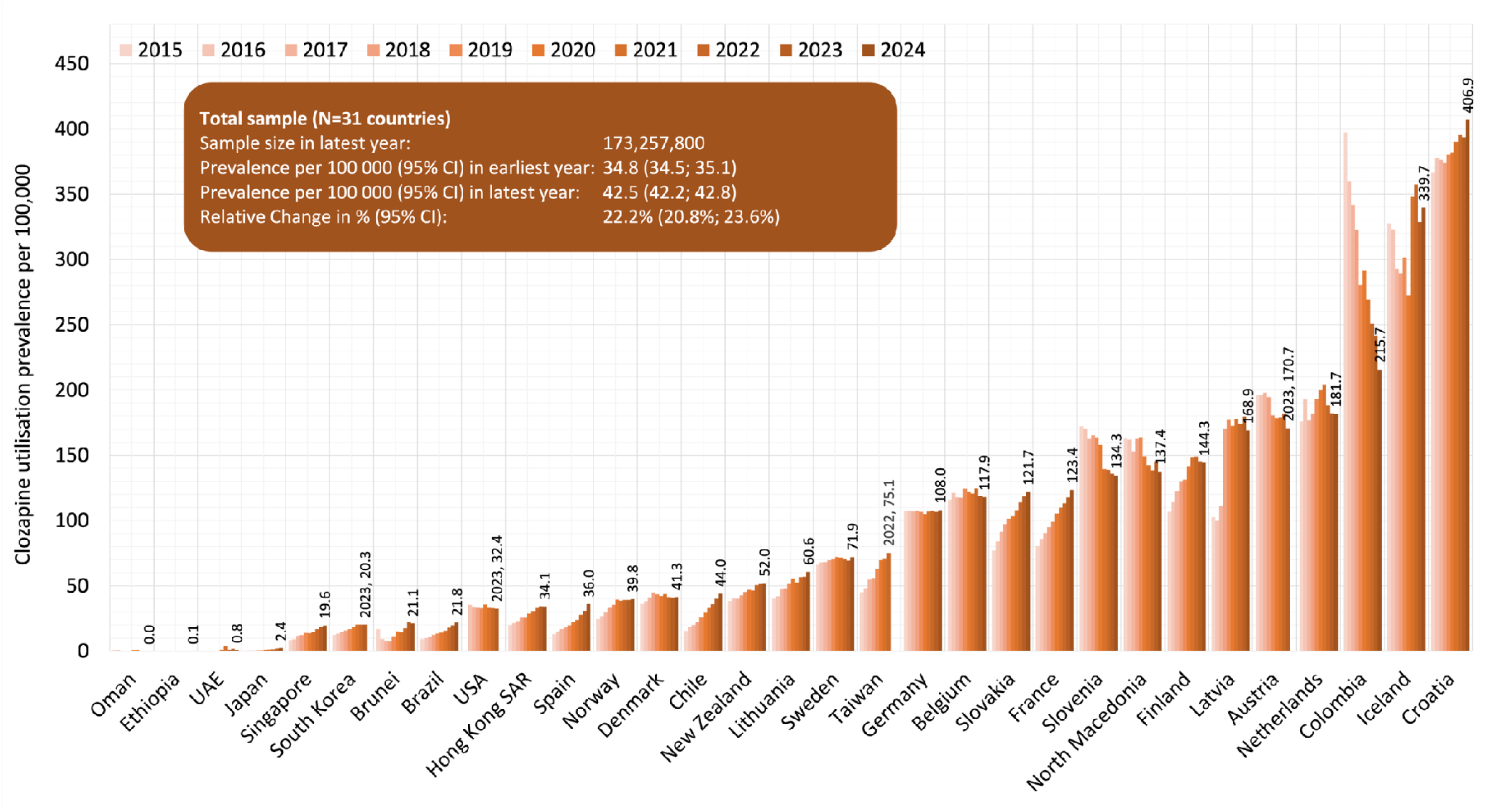
Clozapine utilisation prevalence per 100,000 older adults (≥65 years), 2015–2024. SAR = special administrative region; UAE = United Arab Emirates; USA = United States of America.

### Clozapine use across all countries

Across all countries, clozapine utilisation prevalence in adults increased from 44.7/100,000 (95% CI: [44.5; 44.9]) at the earliest observation to 52.2/100,000 (95% CI: [51.1; 52.4]) at the latest observation within the study period (relative change: +16.9%; 95% CI: [16.3; 17.5]) (**Figure 1**).

### Clozapine use in countries with overall prevalence and consumption estimates only

Among countries that provided overall prevalence estimates only, the prevalence of clozapine use in 2024 ranged from 27.3/100,000 (Argentina) to 314.2/100,000 (Cyprus) (**Table 3**). Between 2015 and 2024, the relative change in prevalence ranged from a decrease of −6.7% (Australia) to an increase of 49.7% (Romania).

**Table 3:**
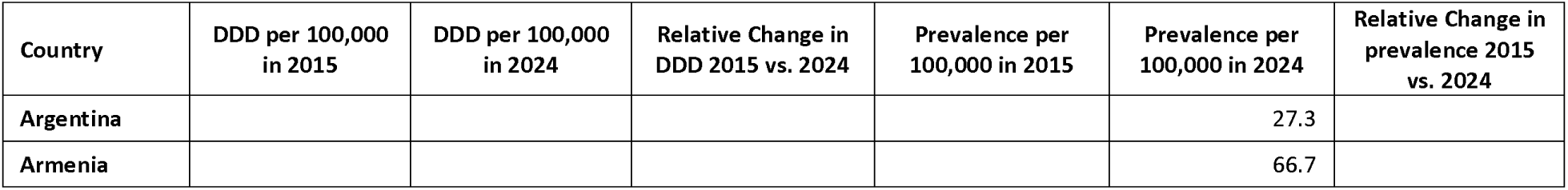
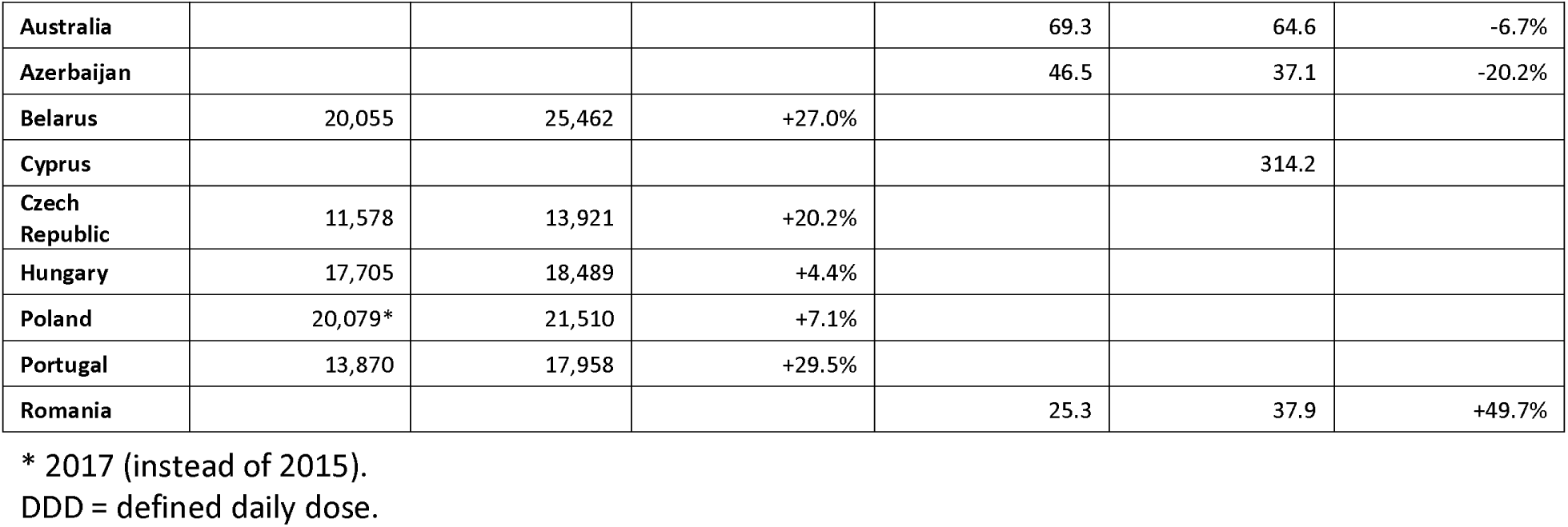
Clozapine utilisation in countries with aggregated data, by country.

Among countries providing consumption data, clozapine use in 2024 ranged from 13,921 DDDs per 100,000 population (Czech Republic) to 25,462 DDDs per 100,000 population (Belarus) (**Table 3**). Over the study period, the relative change in consumption ranged from an increase of 4.4% (Hungary) to 29.5% (Portugal).

## Discussion

### Clozapine use in adults and variation between countries

In this observational study spanning 42 countries/SAR, we observed more than a 1,000-fold difference in clozapine utilisation prevalence between jurisdictions. This variation is substantially greater than the 315-fold variation reported in our previous study, which was predominantly in high-income settings.^20^ The wider disparity observed in the present study likely reflects the inclusion of low- and middle-income countries, where regulatory, infrastructural and workforce barriers may further constrain access to clozapine. Taken together, these findings highlight profound international inequities in access to evidence-based care for TRS and underscore the need for a coordinated global effort to improve access to this highly effective yet underutilised intervention.

Amongst the surveyed countries, Croatia, Colombia and Finland had the highest clozapine utilisation prevalence. In Finland and Croatia, utilisation may partly reflect prescribing beyond TRS, including low-dose clozapine for suicide prevention, although indication data were unavailable to confirm this.^31^ Finland’s high utilisation may also reflect decades of influential clozapine research and successful translation of evidence into clinical practice.^14,17,23^ In Colombia, broader licensing indications for clozapine, historic limited antipsychotic alternatives and the absence of mandatory haematological monitoring up until 2022 may have facilitated greater use. By contrast, the very low utilisation observed in Ethiopia and Oman likely reflects a combination of service centralisation, limited monitoring infrastructure, workforce shortages, geographic barriers and clinician and patient concerns regarding treatment monitoring and adverse effects.^32^

By comparison, our overall findings differ substantially from recent DDD-based analyses of clozapine utilisation derived from pharmaceutical sales data.^33^ Notably, countries that ranked highly in the sales-based study often occupied markedly different positions in our utilisation-prevalence analyses and temporal trends sometimes moved in opposite directions. These discrepancies suggest that sales-based DDD estimates and patient-level utilisation prevalence capture different constructs. From a clinical and public health perspective, prevalence estimates may be more informative because they directly quantify the number of individuals treated and can be compared with disease prevalence to provide a crude estimate of under- or overprescribing.^34^

### Clozapine use by age and sex

In most countries, clozapine utilisation peaked between ages 50 and 59 years. While this partly mirrors the age distribution of schizophrenia^35^, it is likely indicative of delay in the initiation of clozapine, with many individuals historically receiving treatment only after prolonged illness and exposure to non-evidenced based treatment strategies. Interestingly, peak utilisation occurred at younger ages in several Asian and South American countries, potentially suggesting earlier access to clozapine. Sex differences observed in our study likely reflected the higher prevalence of schizophrenia (approximately 1:1.6)^3^ and TRS among men (approximately 1:1.4).^36^ However, peak utilisation occurred approximately 5 to 10 years earlier in men than in women across most countries, consistent with the earlier onset of schizophrenia in males.^37^

### Clozapine use in adolescents and older adults

Several countries, including Colombia, Croatia, and Finland, exceeded our conservative estimates of expected clozapine utilisation among adolescents of 10–20 per 100,000 youths.^20^ This finding is encouraging given the poorer prognosis associated with early-onset schizophrenia and the importance of timely access to effective treatment.^38,39^ Conversely, very low utilisation in younger populations may reflect diagnostic uncertainty, limited clinician familiarity and regulatory restrictions on prescribing in adolescents.

Among older adults, Iceland showed exceptionally high utilisation, particularly among women aged ≥80 years, exceeding more than 800 per 100,000. This may partly reflect clozapine use for Parkinson’s disease psychosis, alongside Iceland’s generally high psychotropic prescribing rates and longer female life expectancy.^40–43^ In contrast, the very high rates observed among middle-aged adults in Croatia are unlikely to be explained by Parkinson’s disease psychosis and may suggest prescribing for additional indications beyond TRS.

### Time trends in clozapine use

A key objective of our study was to examine temporal trends in clozapine use across the surveyed countries. Overall clozapine utilisation prevalence increased by 16.9% during the study period. However, trends varied substantially between countries, ranging from a 71% decrease to a 342% increase. Compared with our previous study, a smaller proportion of countries (52%;16 of 31) demonstrated substantial growth in clozapine utilisation, suggesting that progress has been uneven.^20^ Country-specific policies and service structures likely contributed to these trends. Large increases in Japan and Brazil may reflect expanded access following regulatory and healthcare reforms^20^, whereas France demonstrated sustained long-term growth that may be related to active clinical research networks, professional education and specialised rehabilitation services. In contrast, clozapine utilisation declined in Germany despite longstanding guideline recommendations, highlighting that the drivers of clozapine use remain incompletely understood.^28^ While the overall increase represents a positive development, it remains considerably smaller than the increase in clozapine consumption reported in sales-based analyses (17 vs 39%).^33^

### Potential clozapine underutilisation

International guidelines recommend that all individuals with recognised TRS should be offered clozapine.^1^ Using a conservative estimate of TRS prevalence of approximately 99 per 100,000 population, clozapine utilisation prevalence would ideally approach this level. ^2,3,44^ From our 2024 data, 17 of 31 countries reported clozapine utilisation below this threshold in adults, suggesting potential underutilisation. This treatment gap was notably observed across continents and across countries with differing socioeconomic profiles. Given emerging evidence supporting clozapine use in selected populations beyond classical TRS and proposals advocating earlier use in schizophrenia, the true unmet need may be even greater.^15,45–47^

The reasons for clozapine underutilisation are likely multifactorial. Institutional barriers, including service organisation, insurance coverage, medication costs and access to monitoring infrastructure have been widely described.^48^ Prescriber-related barriers, such as concerns regarding adverse effects, monitoring requirements, ethnic inequalities in prescribing and failure to recognise or act upon TRS, may have also contribute substantially to underuse.^21,49^ Patient-related barriers including concerns regarding blood testing, adverse effects and treatment burden have likewise been described.^50^ Furthermore, country-specific regulations governing clozapine discontinuation and re-exposure following haematological adverse events (that is, “rechallenge”) may further influence utilisation patterns.^7^ Notably, recent guideline updates have increasingly emphasised the central role of clozapine in TRS management to improve patient outcomes. However, evidence suggests that adherence to these recommendations remains suboptimal, indicating that dissemination of evidence alone is insufficient to ensure implementation in clinical practice. ^51–54^

### Association between clozapine utilisation and medico-socio-economic variables

In our exploratory analysis, a notable finding was the absence of any association between clozapine utilisation prevalence and national income, healthcare expenditure, psychiatrist density, urbanicity, mental health expenditure, or monitoring stringency (**Table 3**). These findings suggest that variation in clozapine utilisation may be influenced less by macro-level resources and more by factors such as prescribing culture, clinician attitudes, service organisation, and patient perceptions of treatment. ^50,55^

### Outlook

The persistent underutilisation of clozapine, despite its well-established benefits in symptom control, mortality reduction and cost-effectiveness,^18^ represents a long-standing global conundrum in the treatment of schizophrenia. This paradox is particularly striking given clozapine’s comparatively favourable safety profile^56^ and the fact that its potential to save lives substantially outweighs the risk of rare but serious adverse events.^57^ Nonetheless, clozapine remains underused across many health systems, constituting a major and enduring public health challenge.

Although the removal or relaxation of haematological monitoring requirements has generated optimism that clozapine use might increase, both in the United States^58^ and Europe,^59^ such regulatory changes alone appear insufficient to drive sustained improvements in utilisation.^60^ The persistence of clozapine underutilisation over several decades raises critical questions regarding which interventions have been attempted to increase uptake, which have been effective and scalable and which barriers have impeded their broader implementation across countries. Evidence increasingly indicates that sustainable improvements require coordinated, multi-level interventions. These may include dedicated clozapine services, streamlined monitoring pathways, audit-and-feedback systems, integrated physical and mental healthcare, specialist consultation networks, electronic identification of eligible patients and strong local clinical leadership. ^21,61,62,63–65^ Educational initiatives remain important but are unlikely to be sufficient in isolation.

Given substantial variation in healthcare systems internationally, effective solutions are unlikely to be universal. Instead, implementation strategies tailored to local barriers and informed by implementation science will be required. Improving access to clozapine represents a major opportunity to reduce the burden of TRS and may provide a broader model for improving access to other effective but underutilised psychotropic treatments.

The major strengths of this study include its unique scale, encompassing individual-level data from more than one billion people across multiple continents, the use of nationally representative prescription and dispensing datasets and the availability of age- and sex-specific analyses. The 10-year observation period also enabled robust assessment of temporal trends.

Notwithstanding, several limitations should be acknowledged. Regional rather than national data were available for some countries, and age- and sex-stratified information was unavailable in all settings. Differences in healthcare systems and data sources may also affect comparability. For a small number of countries, DDD-based estimates were used when individual-level data were unavailable. In addition, outpatient prescription data do not capture individuals residing long-term in psychiatric hospitals or forensic institutions, potentially leading to modest underestimation of utilisation prevalence. Despite extensive international coverage, countries were included on the basis of data availability rather than a predefined sampling strategy. Consequently, findings may not fully represent global clozapine utilisation patterns, and utilisation may be overestimated if countries with lower prescribing rates were underrepresented. The ecological analyses should be interpreted cautiously because they were exploratory and not designed for causal inference. Finally, information on treatment indication, dosage, and incidence of clozapine prescribing was unavailable, limiting our ability to distinguish clozapine use for TRS from use for other indications.

## Conclusion

International variation in clozapine utilisation is substantial, with more than a 1,000-fold difference observed between countries. Although clozapine utilisation increased overall during the study period, utilisation remained below expected treatment need in many settings. The concentration of clozapine use among individuals aged 50 to 59 years further suggests continuing delays in treatment initiation. The absence of associations with a range of socioeconomic and healthcare-system indicators suggests that utilisation is driven less by national resources and more by factors such as service organisation, prescribing culture, and clinician and patient attitudes. Future research should prioritise the development and evaluation of targeted implementation strategies to improve timely and equitable access to clozapine.

## Statements

## Acknowledgement

The lead authors would like to thank those colleagues all over the world who helped with identifying data sources for our study.

Japan: The authors from Japan were supported by Novartis Pharma in the acquisition of the CPMS data in Japan. Spain: This study was conducted using anonymized data provided by the Agency for Quality and Assessment of Catalonia (Agència de Qualitat i Avaluació Sanitàries de Catalunya; AQuAS), within the framework of the Data Analytics Program for Health Research and Innovation (Programa d’Analítica de Dades per a la Recerca i la Innovació en Salut; PADRIS) Programme.

Taiwan: Professor Cynthia Wei-Sheng Lee, and Chia-Hsun Chung for Taiwan data collection and inspection.

## Conflicts of interest

Christian J Bachmann is a member of the European Clozapine Task Force, an informal, not-for-profit association of European clinicians with an interest in improving access to clozapine for patients with schizophrenia.

Robert Bittner has received advisory board fees from Newron and speaker fees from Recordati Pharma GmbH. Member of the European Clozapine Task Force.

Andreja Celofiga participated in lectures for Krka, Gedeon Richter, Bonifar, Lundbeck, Viatris, Teva, and Janssen; and participated in a clinical trial for Krka. Member of the European Clozapine Task Force.

Robert O. Cotes has received research funding from Otsuka, Boehringer Ingelheim, Karuna, and Alkermes; served as a consultant to the Clozapine Product Manufacturers Group, Boehringer Ingelheim, Saladax Biomedical, and IQVIA/Cronos.

Vlad Dionisie is on speakers bureaus for Johnson & Johnson Romania SRL, Lundbeck Romania SRL. Member of the European Clozapine Task Force.

Ary Gadelha: Honoraria and consultant/advisor from Daiichi-Sankyo, Adium, Johnson & Johnson, Critália, Lundbeck, Ache, and Boehringer Ingelheim.

Daniel Guinart has been a consultant and/or advisor or has received honoraria from: Angelini, Otsuka, Lundbeck, Teva and Viatris. Member of the European Clozapine Task Force.

Jimmy Lee has received honoraria, served as a consultant or advisory board member from Otsuka, Janssen,

Lundbeck, Sumitomo Pharmaceuticals, Boehringer Ingelheim, ThoughtFull World Pte. Ltd. and Singapore Deep-Tech Alliance.

Raffael Massuda has received honoraria, served as a consultant or advisory board member from Adium, Johnson & Johnson, Daichii Sankyo, and Boehringer Ingelheim.

Claude Mawa received funding to visit conferences from Angelini Pharma, Neroxpharm, Adamed, Berlin Chemia, Biopharm, Plus pharma, Aurovitas Pharma, Pro.med.pl

Cristian Mena is an advisor to the Department of Mental Health (DIPRECE) at the Chilean Ministry of Health, involved in initiatives to improve access to clozapine for people with schizophrenia in Chile (e.g., clinical guidelines and electronic monitoring systems).

Uladzimir Pikirenia: Received funding to visit conference from Plus Pharma, received scholarship from Tobacco Harm Reduction Scholarship Program.

Maria Gabriela Puiu: Speakers bureaus for Johnson & Johnson Romania SRL, Lundbeck Romania SRL, MERCK SHARP & DOHME Romania SRL, Angelini Pharmaceuticals Romania SRL, TERAPIA SA, SC KRKA Romania SRL, SERVIER PHARMA S.R.L., SC AstraZeneca Pharma SRL, IQVIA RDS Eastern Holdings GmbH.

Julieta Ramirez has been a consultant and/or advisor or has received honoraria from: Acadia, Adium, Bago, Baliarda, Bristol-Meyers-Squibb, Boehringer-Ingelheim, Casasco, Elea, Gador, Gedeon Richter, Janssen/J&J, Lundbeck, Megalabs, Nutricia Bagó-Danone, Raffo, Roemmers, Siegfried, Teva.

Rifai Farid and Fariza Sani work in the psychiatry service that is evaluated in this paper. The contributions in this paper are their own and do not necessarily represent the Ministry of Health, Brunei Darussalam.

Dan Siskind serves on the Viatris Australian Clozapine Quality Advisory Committee and DS has received honorarium for independent educational talks from Servier, Viatris and Lundbeck.

Heidi Taipale has received grants from Janssen paid to employer institution and lecture fees from Gedeon Richter, Janssen, Lundbeck, and Otsuka, outside of the submitted work.

David Taylor received honoraria, served as a consultant or advisory board member for Idorsia, Otsuka, Janssen, Lundbeck, and Viatris.

Hélène Verdoux is a member of the European Clozapine Task Force.

The remaining authors report no competing interests.

## Data availability statement

The aggregated data underlying the results reported in this study are available from the corresponding author upon reasonable request, subject to approval by the site investigators and their respective data providers.

## SUPPLEMENTARY MATERIAL

**Figure S1:**
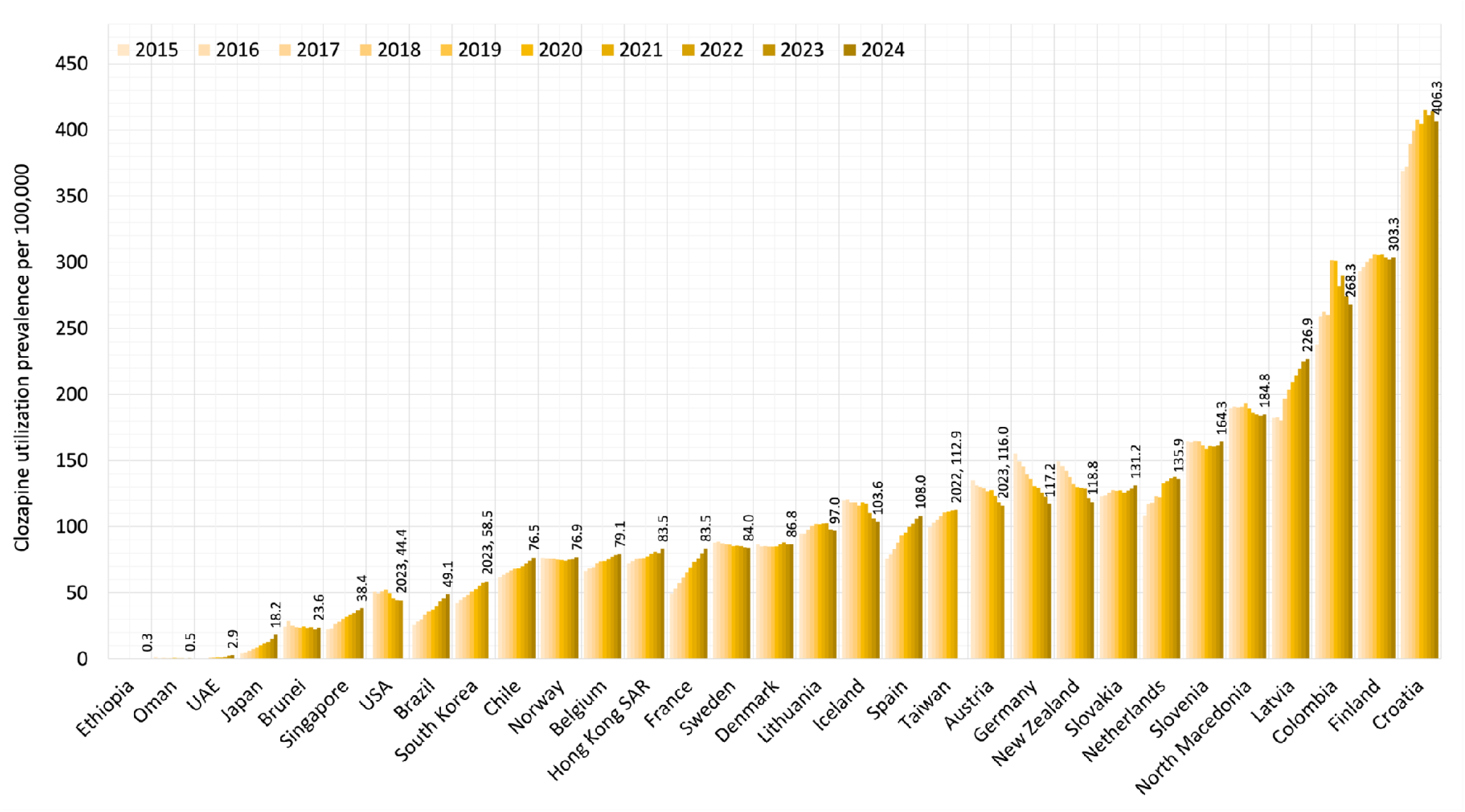
Age- and sex-standardised clozapine utilisation prevalence per 100,000 adults (20–64 years), 2015–2024. SAR = special administrative region; UAE = United Arab Emirates; USA = United States of America.

**Figure S2:**
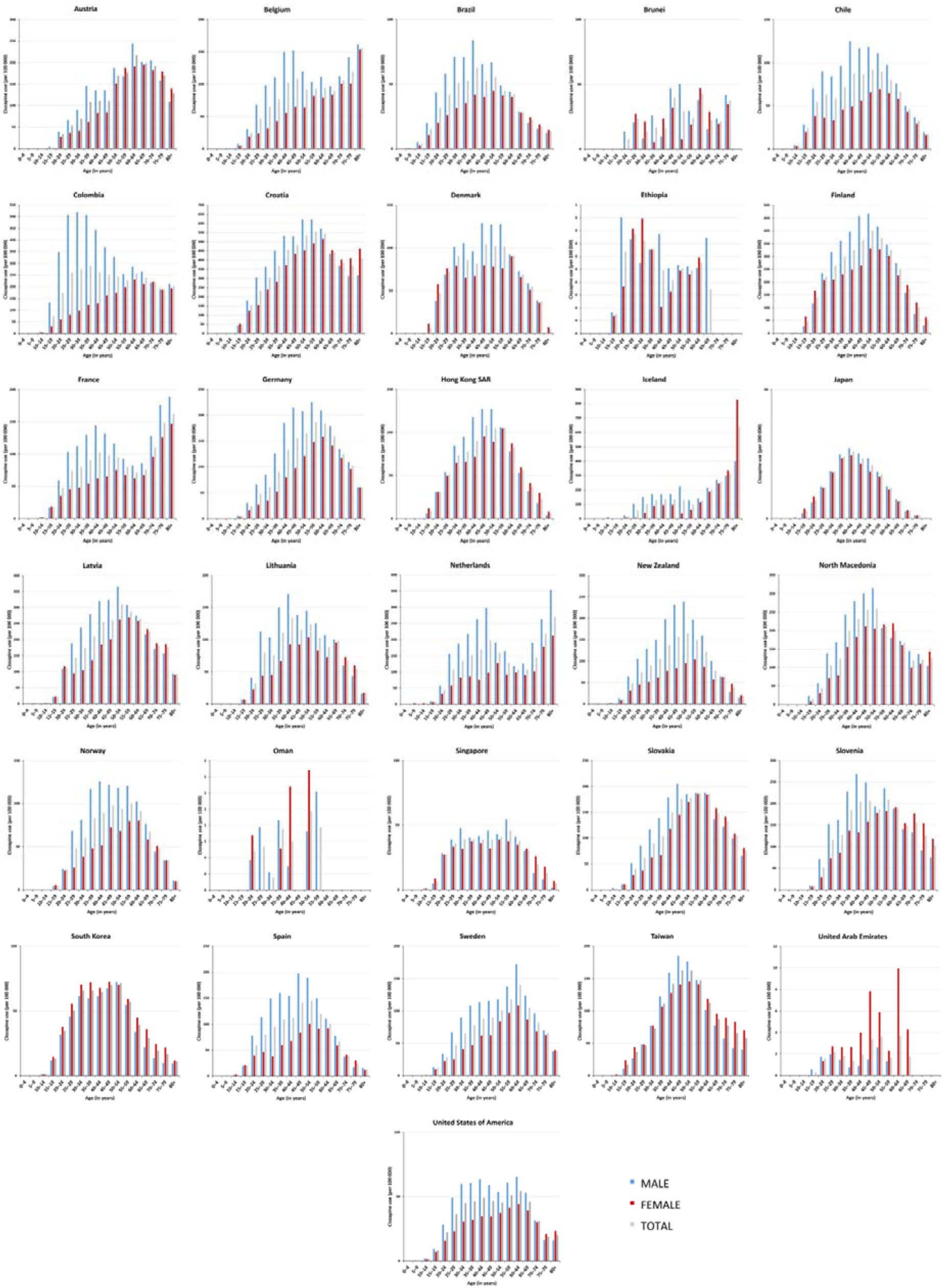
Clozapine utilisation prevalence per 100,000 persons in 2024 (or most recent year available) per country, by sex and age.

**Table S1:**
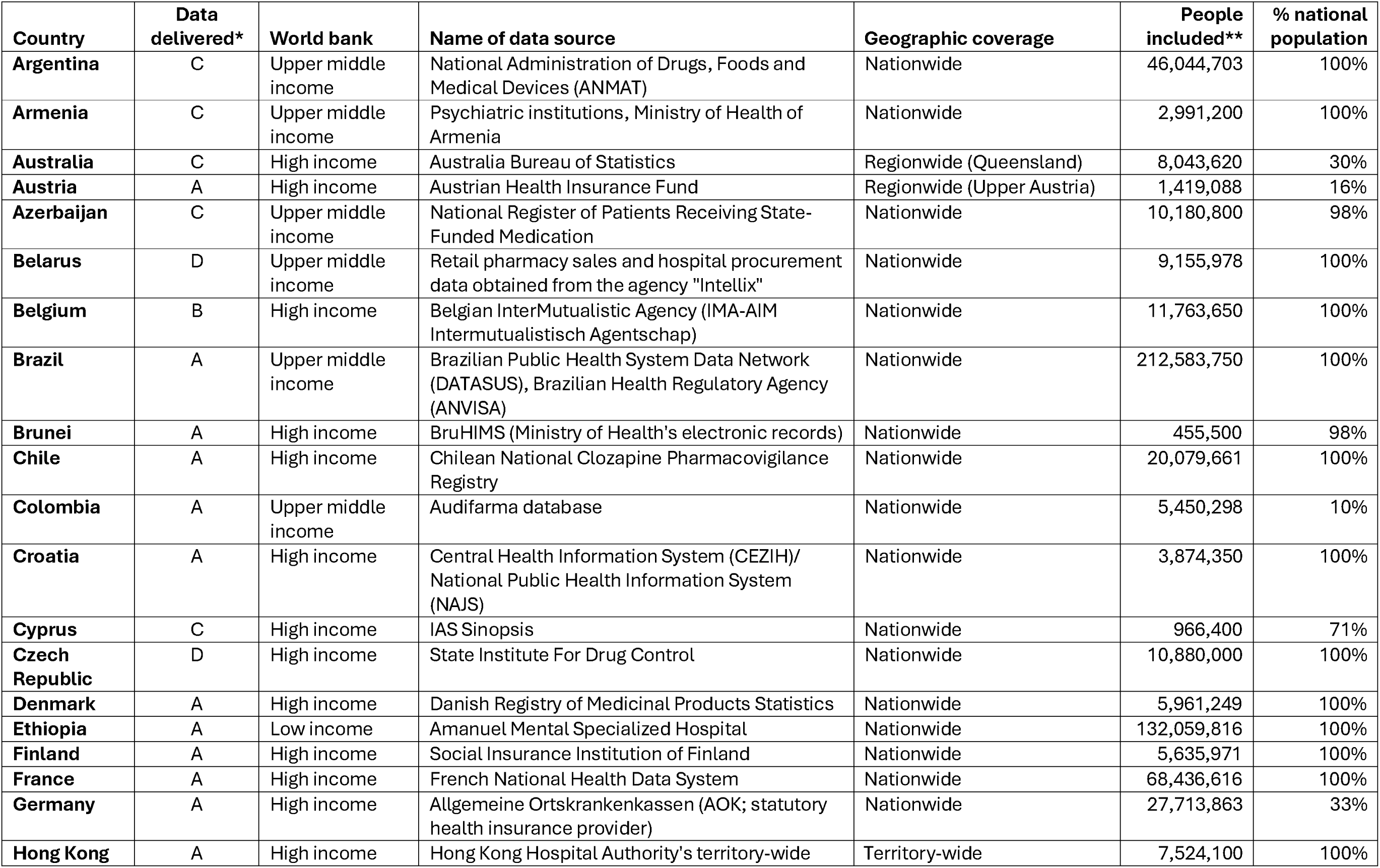
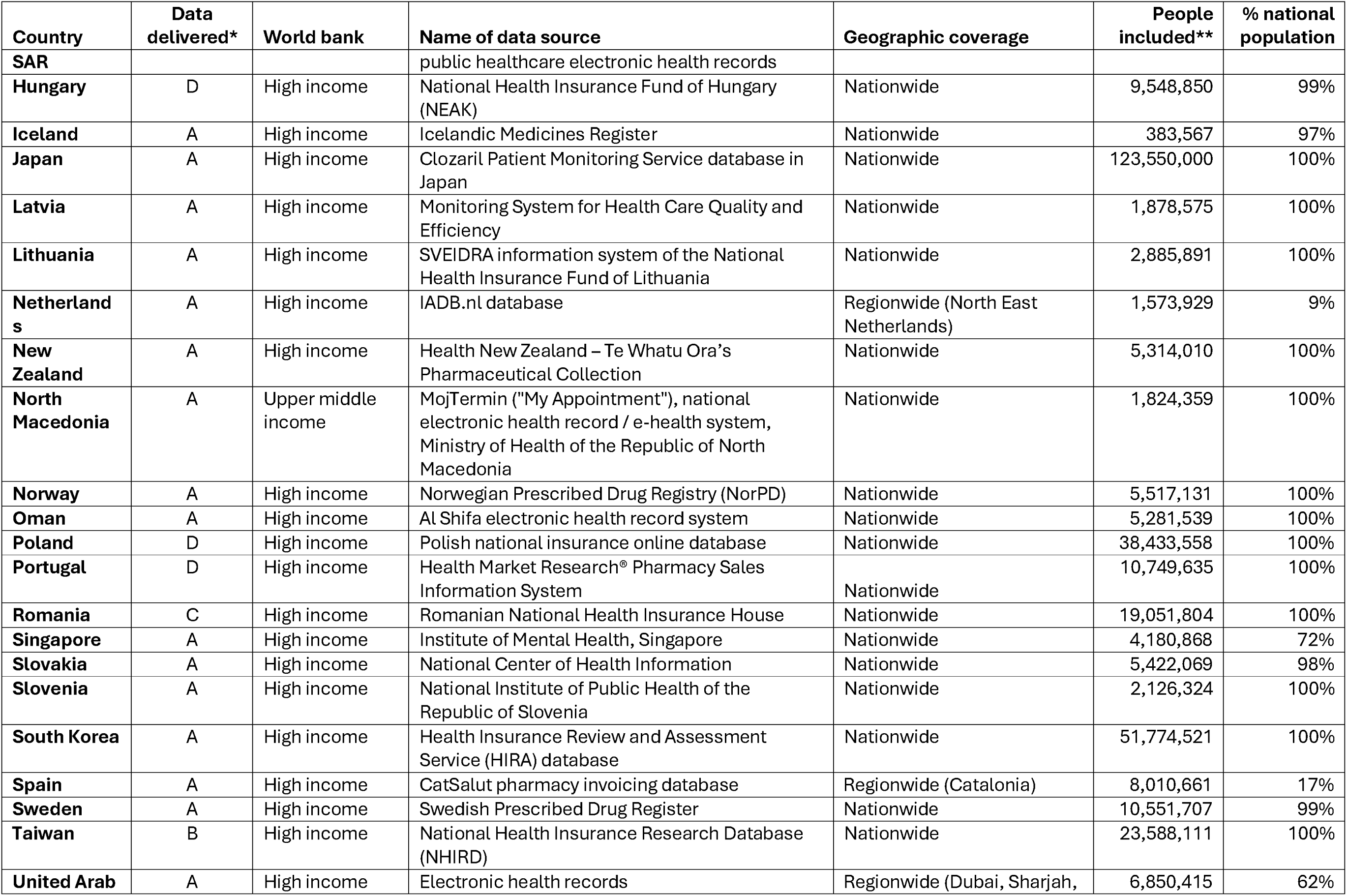
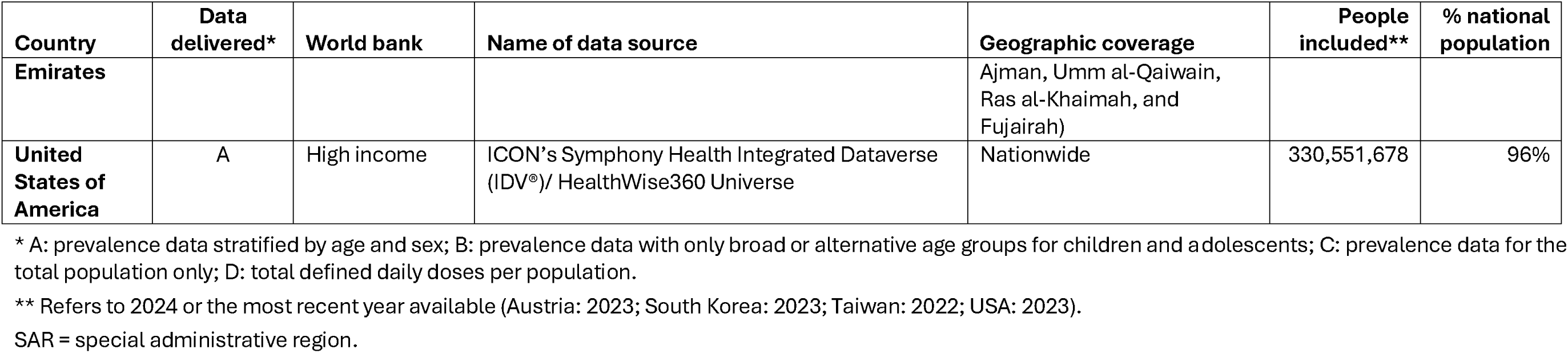
Data source characteristics, by country.

| **Country** | **Data delivered\*** | **World bank** | **Name of data source** | **Geographic coverage** | **People included\*\*** | **% national population** |
| --- | --- | --- | --- | --- | --- | --- |
| **Argentina** | C | Upper middle income | National Administration of Drugs, Foods and Medical Devices (ANMAT) | Nationwide | 46,044,703 | 100% |
| **Armenia** | C | Upper middle income | Psychiatric institutions, Ministry of Health of Armenia | Nationwide | 2,991,200 | 100% |
| **Australia** | C | High income | Australia Bureau of Statistics | Regionwide (Queensland) | 8,043,620 | 30% |
| **Austria** | A | High income | Austrian Health Insurance Fund | Regionwide (Upper Austria) | 1,419,088 | 16% |
| **Azerbaijan** | C | Upper middle income | National Register of Patients Receiving State-Funded Medication | Nationwide | 10,180,800 | 98% |
| **Belarus** | D | Upper middle income | Retail pharmacy sales and hospital procurement data obtained from the agency "Intellix" | Nationwide | 9,155,978 | 100% |
| **Belgium** | B | High income | Belgian InterMutualistic Agency (IMA-AIM Intermutualistisch Agentschap) | Nationwide | 11,763,650 | 100% |
| **Brazil** | A | Upper middle income | Brazilian Public Health System Data Network (DATASUS), Brazilian Health Regulatory Agency (ANVISA) | Nationwide | 212,583,750 | 100% |
| **Brunei** | A | High income | BruHIMS (Ministry of Health's electronic records) | Nationwide | 455,500 | 98% |
| **Chile** | A | High income | Chilean National Clozapine Pharmacovigilance Registry | Nationwide | 20,079,661 | 100% |
| **Colombia** | A | Upper middle income | Audifarma database | Nationwide | 5,450,298 | 10% |
| **Croatia** | A | High income | Central Health Information System (CEZIH)/ National Public Health Information System (NAJS) | Nationwide | 3,874,350 | 100% |
| **Cyprus** | C | High income | IAS Synopsis | Nationwide | 966,400 | 71% |
| **Czech Republic** | D | High income | State Institute For Drug Control | Nationwide | 10,880,000 | 100% |
| **Denmark** | A | High income | Danish Registry of Medicinal Products Statistics | Nationwide | 5,961,249 | 100% |
| **Ethiopia** | A | Low income | Amanuel Mental Specialized Hospital | Nationwide | 132,059,816 | 100% |
| **Finland** | A | High income | Social Insurance Institution of Finland | Nationwide | 5,635,971 | 100% |
| **France** | A | High income | French National Health Data System | Nationwide | 68,436,616 | 100% |
| **Germany** | A | High income | Allgemeine Ortskrankenkassen (AOK; statutory health insurance provider) | Nationwide | 27,713,863 | 33% |
| **Hong Kong** | A | High income | Hong Kong Hospital Authority's territory-wide | Territory-wide | 7,524,100 | 100% |

| Country | Data delivered* | World bank | Name of data source | Geographic coverage | People included** | % national population |
| --- | --- | --- | --- | --- | --- | --- |
| **SAR** |  |  | public healthcare electronic health records |  |  |  |
| **Hungary** | D | High income | National Health Insurance Fund of Hungary (NEAK) | Nationwide | 9,548,850 | 99% |
| **Iceland** | A | High income | Icelandic Medicines Register | Nationwide | 383,567 | 97% |
| **Japan** | A | High income | Clozaril Patient Monitoring Service database in Japan | Nationwide | 123,550,000 | 100% |
| **Latvia** | A | High income | Monitoring System for Health Care Quality and Efficiency | Nationwide | 1,878,575 | 100% |
| **Lithuania** | A | High income | SVEIDRA information system of the National Health Insurance Fund of Lithuania | Nationwide | 2,885,891 | 100% |
| **Netherlands** | A | High income | IADB.nl database | Regionwide (North East Netherlands) | 1,573,929 | 9% |
| **New Zealand** | A | High income | Health New Zealand – Te Whatu Ora’s Pharmaceutical Collection | Nationwide | 5,314,010 | 100% |
| **North Macedonia** | A | Upper middle income | MojTermin ("My Appointment"), national electronic health record / e-health system, Ministry of Health of the Republic of North Macedonia | Nationwide | 1,824,359 | 100% |
| **Norway** | A | High income | Norwegian Prescribed Drug Registry (NorPD) | Nationwide | 5,517,131 | 100% |
| **Oman** | A | High income | Al Shifa electronic health record system | Nationwide | 5,281,539 | 100% |
| **Poland** | D | High income | Polish national insurance online database | Nationwide | 38,433,558 | 100% |
| **Portugal** | D | High income | Health Market Research® Pharmacy Sales Information System | Nationwide | 10,749,635 | 100% |
| **Romania** | C | High income | Romanian National Health Insurance House | Nationwide | 19,051,804 | 100% |
| **Singapore** | A | High income | Institute of Mental Health, Singapore | Nationwide | 4,180,868 | 72% |
| **Slovakia** | A | High income | National Center of Health Information | Nationwide | 5,422,069 | 98% |
| **Slovenia** | A | High income | National Institute of Public Health of the Republic of Slovenia | Nationwide | 2,126,324 | 100% |
| **South Korea** | A | High income | Health Insurance Review and Assessment Service (HIRA) database | Nationwide | 51,774,521 | 100% |
| **Spain** | A | High income | CatSalut pharmacy invoicing database | Regionwide (Catalonia) | 8,010,661 | 17% |
| **Sweden** | A | High income | Swedish Prescribed Drug Register | Nationwide | 10,551,707 | 99% |
| **Taiwan** | B | High income | National Health Insurance Research Database (NHIRD) | Nationwide | 23,588,111 | 100% |
| **United Arab** | A | High income | Electronic health records | Regionwide (Dubai, Sharjah, | 6,850,415 | 62% |
| **Emirates** |  |  |  | Ajman, Umm al-Qaiwain, Ras al-Khaimah, and Fujairah) |  |  |
| **United States of America** | A | High income | ICON's Symphony Health Integrated Dataverse (IDV®)/ HealthWise360 Universe | Nationwide | 330,551,678 | 96% |

\* A: prevalence data stratified by age and sex; B: prevalence data with only broad or alternative age groups for children and adolescents; C: prevalence data for the total population only; D: total defined daily doses per population.
\*\* Refers to 2024 or the most recent year available (Austria: 2023; South Korea: 2023; Taiwan: 2022; USA: 2023).
SAR = special administrative region.

**Table S2:** Ethical approval procedures, by country.

| **Country/region** | **Ethical approval procedure** |
| --- | --- |
| **Argentina** | No ethical approval was required because the study used anonymized, aggregated secondary data obtained from the National Administration of Drugs, Foods and Medical Devices (ANMAT). No individual-level patient information was accessed and no human subjects were directly involved in the research. |
| **Armenia** | Data were obtained from annual reports of the main psychiatric clinics in Armenia. |
| **Australia** | No ethical approval was necessary as these data were publicly available. |
| **Austria** | Ethical approval was not required as the pseudonymised data was provided for research purposes under a contract with the Austrian Health Insurance Fund. |
| **Azerbaijan** | No ethical approval was necessary as these data were publicly available. |
| **Belarus** | No ethical approval was required because the study used anonymized, aggregated secondary data. |
| **Belgium** | No ethical approval was necessary as these data were fully anonymous and based on governmental medication reimbursement records |
| **Brazil** | No ethical approval was necessary as these data were publicly available. |
| **Brunei** | Ethical approval was sought from formally from the Biomedical Research & Ethics Unit, Ministry of Health, Brunei Darussalam |
| **Chile** | Data were obtained as aggregated, anonymised statistics through a formal public transparency request under Chilean Law 20.285 on Access to Public Information. No individual-level data were used. According to national regulations, ethical approval was not required for this type of secondary use of aggregated data. |
| **Colombia** | According to the respective national regulations, no ethical approval was necessary for this study |
| **Croatia** | No ethical approval was needed, as the study is based on secondary analysis of aggregated data, which do not allow identification of individual participants |
| **Cyprus** | Ethical approval not required due to anonymized, secondary data |
| **Czech Republic** | No ethical approval was needed, as the study is based on secondary analysis of aggregated data, which do not allow identification of individual participants |
| **Denmark** | According to the respective national regulations, no ethical approval was necessary for this study |
| **Ethiopia** | The ethical review board of Amanuel Mental Specialised Hospital approved the study (ref. Am/146/4/7) |
| **Finland** | According to the respective national regulations, no ethical approval was necessary for this study |
| **France** | In accordance with the permanent regulatory access to the database granted to Caisse National de l'Assurance Maladie (Cnam), this study did not require specific authorization from the French Protection Authority. |
| **Germany** | According to the respective national regulations, no ethical approval was necessary for this study |
| **Hong Kong SAR** | The Central Institutional Review Board of the Hospital Authority approved this study (Ref: CIRB-2022-015-5). The requirement for informed consent was waived because the study used only anonymised electronic health records |
| **Hungary** | No ethical approval was necessary as these data were publically available |
| **Iceland** | All data used in this study consisted solely of anonymized patient records, and no results were reported for estimates involving ten patients or fewer |
| **Japan** | This study was approved by the Ethics Committee of the Tohoku University Graduate School of Medicine (Approval ID: 2023-1-856/857) |
| **Latvia** | According to local data protection regulations, no approval from the ethics committee was required for this study |
| **Lithuania** | No ethical approval was necessary for this study because the data is aggregated and similar analyses are part of the functions of the National Health Insurance Fund |
| **Netherlands** | According to the respective national regulations, no ethical approval was necessary for this study |
| **New Zealand** | Biomedical Sciences Research Ethics Committee, University of Bath (Ref. 11860-14627) |
| **North Macedonia** | No ethical approval was necessary as the data were anonymised/aggregated routine data provided by the Ministry of Health. |
| **Norway** | According to the respective national regulations in Norway, no ethical approval was necessary for this study. |
| **Oman** | Ethical approval for the study was granted by the Research and Ethical Committee of the Ministry of Health in Muscat, Oman (Ref. MH/DGHS/DPT/369/2025) |
| **Poland** | No ethical approval was required for this study because only anonymised, aggregated data from publicly available national sources were used. |
| **Portugal** | No ethical approval was necessary for this study as anonymised aggregate data was used. |
| **Romania** | No ethical approval was necessary as these data were categorized as information of public interest and were publicly available by request according to the national law 554/2001. |
| **Singapore** | The Institutional review committee waived requirements for full ethics review of the present analyses (IRRC-RNR-2025-003) because only anonymous data was used for the present study. |
| **Slovakia** | No approval necessary, as anonymised secondary data were used |
| **Slovenia** | The study was approved by the Medical Ethics Committee of the University Medical Center Maribor (reference UKC-MB-KME-4/26) |
| **South Korea** | The Institutional review committee waived requirements for full ethics review of the present analyses (S2024-2031) because only anonymous data was used for the present study. |
| **Spain** | No ethics approval was necessary |
| **Sweden** | No ethical approval was necessary as these data were publicly available. |
| **Taiwan** | The study was approved by the Institutional Review Board of China Medical University Hospital (approval number: CMUH112-REC1-117(CR-2)) |
| **United Arab Emirates** | Ethical approval for this retrospective study was obtained from the Ministry of Health and Prevention Research Ethics Committee, United Arab Emirates (MOHAP/DXB-REC/M.J.J/No. 91/2024). |
| **United States of America** | Ethical approval was not required study because the analysis used de-identified, aggregate data only. The investigators did not have access to protected health information, individual-level identifiable data, or any re-identification key, and there was no interaction or intervention with human participants. A non-human subjects research form from Emory University is available upon request. |

1 SAR = special administrative region.

**Table S3:**
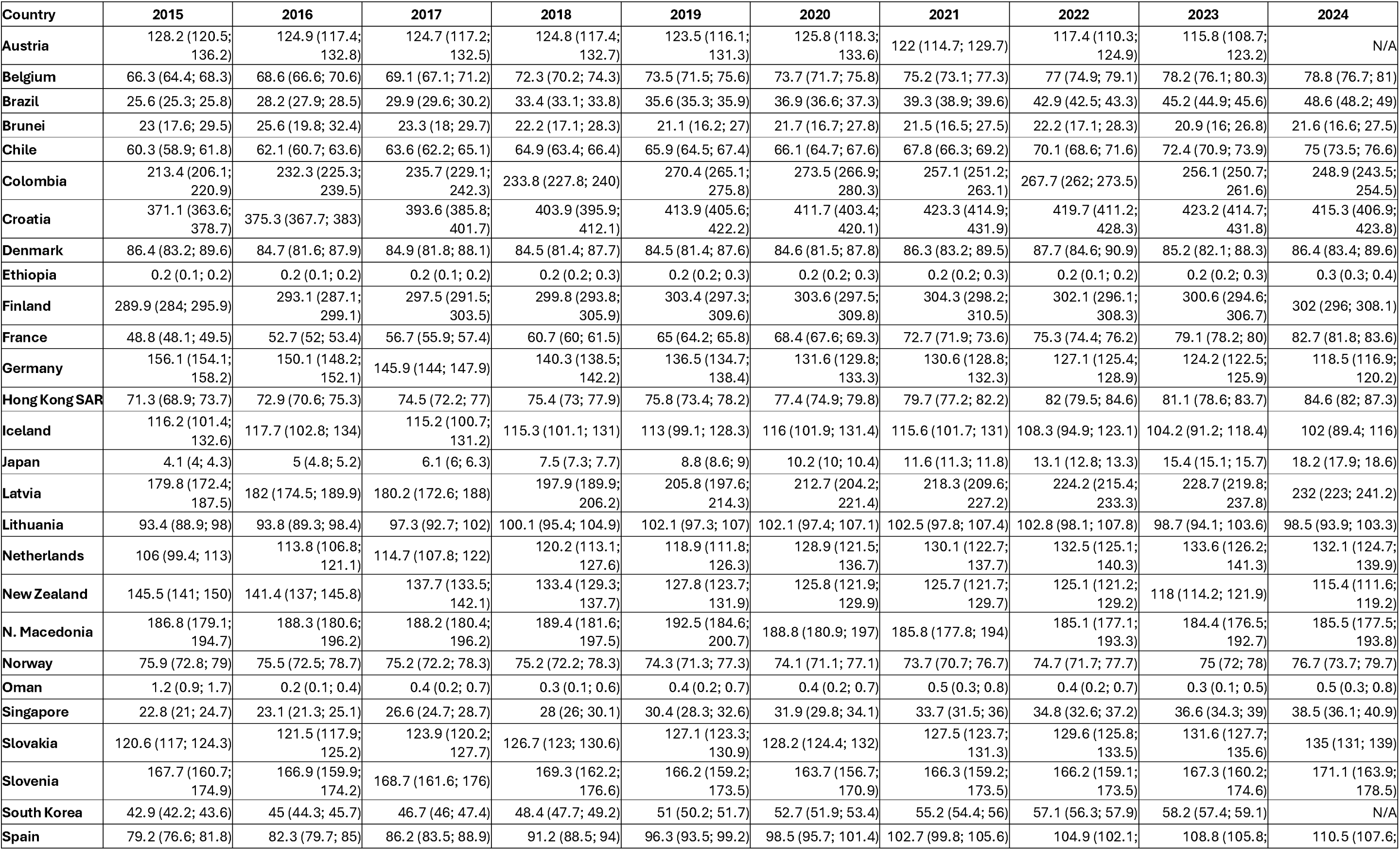
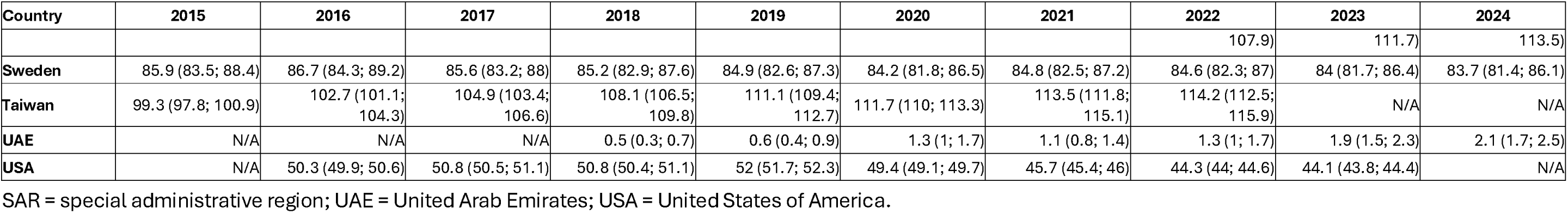
Clozapine utilisation prevalence (and 95% confidence intervals) per 100,000 adults (20–64 years), 2015–2024.

**Table S4:** Clozapine utilisation prevalence (and 95% confidence intervals) per 100,000 adolescents (10–19 years), 2015–2024.

| Country | 2015 | 2016 | 2017 | 2018 | 2019 | 2020 | 2021 | 2022 | 2023 | 2024 |
| --- | --- | --- | --- | --- | --- | --- | --- | --- | --- | --- |
| Austria | 5 (2.2; 9.9) | 7 (3.5; 12.5) | 8.4 (4.4; 14.3) | 9.7 (5.4; 15.9) | 5.8 (2.6; 10.9) | 5.2 (2.2; 10.2) | 3.2 (1; 7.5) | 2.5 (0.7; 6.4) | 1.2 (0.2; 4.5) | N/A |
| Brazil | 3.5 (3.3; 3.7) | 4.1 (3.9; 4.3) | 4.5 (4.3; 4.7) | 5.3 (5; 5.5) | 5.9 (5.7; 6.2) | 6.1 (5.8; 6.4) | 6.8 (6.5; 7.1) | 8 (7.7; 8.3) | 9.1 (8.8; 9.5) | 9.8 (9.4; 10.1) |
| Brunei | 1.4 (0; 8) | 1.4 (0; 7.9) | 1.6 (0; 8.9) | 0 (0; 5.9) | 0 (0; 5.7) | 1.6 (0; 8.8) | 0 (0; 5.7) | 0 (0; 5.8) | 0 (0; 5.9) | 0 (0; 6.1) |
| Chile | 6.3 (5.3; 7.3) | 7.2 (6.2; 8.3) | 8.4 (7.3; 9.7) | 9.9 (8.7; 11.2) | 11.8 (10.5; 13.3) | 11.8 (10.5; 13.2) | 11.5 (10.2; 12.9) | 11.5 (10.3; 12.9) | 13.4 (12; 14.9) | 13.7 (12.3; 15.2) |
| Colombia | 86 (74.9; 98.2) | 95.8 (85.3; 107.2) | 99.7 (90; 110.1) | 85.5 (77.2; 94.4) | 97.7 (90; 105.8) | 94.5 (85.1; 104.8) | 89.1 (80.9; 97.9) | 73.4 (66.5; 80.8) | 62.4 (56.3; 68.9) | 42.7 (37.6; 48.3) |
| Croatia | 18.1 (14.3; 22.6) | 18.2 (14.3; 22.8) | 21.2 (16.9; 26.2) | 20.4 (16.2; 25.4) | 23.2 (18.6; 28.5) | 19.7 (15.5; 24.7) | 22.4 (17.9; 27.6) | 24.4 (19.7; 29.8) | 20.5 (16.2; 25.5) | 24.4 (19.7; 29.9) |
| Denmark | 2.8 (1.7; 4.3) | 4.1 (2.7; 5.9) | 3.4 (2.1; 5) | 1.9 (1; 3.3) | 1.5 (0.7; 2.7) | 2.2 (1.2; 3.6) | 2.5 (1.5; 4) | 4.4 (3; 6.3) | 3.5 (2.3; 5.3) | 2.8 (1.7; 4.4) |
| Ethiopia | 0 (0; 0) | 0 (0; 0.1) | 0 (0; 0.1) | 0 (0; 0) | 0 (0; 0.1) | 0 (0; 0.1) | 0 (0; 0) | 0 (0; 0) | 0 (0; 0.1) | 0 (0; 0.1) |
| Finland | 25 (21.2; 29.4) | 23.1 (19.4; 27.3) | 25.6 (21.7; 30) | 23.4 (19.7; 27.6) | 23 (19.3; 27.1) | 24.4 (20.6; 28.6) | 24.3 (20.6; 28.5) | 23.1 (19.5; 27.2) | 24.2 (20.5; 28.3) | 24.1 (20.5; 28.2) |
| France | 3.8 (3.4; 4.2) | 4.2 (3.8; 4.7) | 4.6 (4.2; 5.1) | 5.6 (5.1; 6.1) | 6.3 (5.8; 6.8) | 7 (6.5; 7.6) | 8 (7.4; 8.6) | 8.8 (8.1; 9.4) | 8.9 (8.3; 9.6) | 10 (9.3; 10.7) |
| Germany | 4 (3.2; 4.9) | 3.9 (3.2; 4.8) | 3.6 (2.9; 4.4) | 3.8 (3.1; 4.6) | 3.9 (3.1; 4.7) | 3.3 (2.6; 4.1) | 3.2 (2.6; 4) | 3.4 (2.8; 4.2) | 3.2 (2.5; 3.9) | 2.7 (2.1; 3.3) |
| Hong Kong SAR | 5.7 (4; 7.9) | 5.8 (4.1; 8.1) | 5.6 (3.8; 7.8) | 5.6 (3.8; 7.9) | 5 (3.3; 7.2) | 5.6 (3.8; 7.9) | 4.7 (3.1; 6.9) | 6.8 (4.8; 9.3) | 5 (3.4; 7.1) | 4.4 (2.9; 6.5) |
| Iceland | 2.3 (0.1; 12.9) | 2.3 (0.1; 12.9) | 0 (0; 8.5) | 0 (0; 8.4) | 0 (0; 8.3) | 0 (0; 8.1) | 0 (0; 8) | 2.1 (0.1; 11.8) | 2.1 (0.1; 11.5) | 2 (0.1; 11.3) |
| Japan | 0.3 (0.2; 0.4) | 0.4 (0.3; 0.5) | 0.5 (0.3; 0.6) | 0.5 (0.4; 0.7) | 0.5 (0.4; 0.7) | 0.6 (0.4; 0.7) | 0.6 (0.5; 0.8) | 0.8 (0.6; 1) | 1.4 (1.2; 1.7) | 1.6 (1.4; 1.9) |
| Latvia | 9.5 (5.5; 15.1) | 13.4 (8.6; 19.9) | 7.7 (4.2; 13) | 11.4 (7; 17.4) | 11.6 (7.3; 17.6) | 12 (7.6; 18) | 16.1 (11; 22.9) | 16.6 (11.4; 23.4) | 11.2 (7; 16.9) | 12.1 (7.8; 18) |
| Lithuania | 3.9 (2; 6.8) | 3.7 (1.9; 6.7) | 8.1 (5.1; 12.2) | 7.6 (4.7; 11.7) | 5.6 (3.1; 9.2) | 6.7 (4; 10.6) | 5.5 (3.1; 9.1) | 5.1 (2.8; 8.6) | 4.2 (2.2; 7.4) | 3.5 (1.7; 6.4) |
| Netherlands | 10.4 (6.2; 16.2) | 8.4 (4.7; 13.9) | 6.1 (3; 10.9) | 6.6 (3.4; 11.5) | 3.9 (1.6; 8.1) | 5.3 (2.4; 10.1) | 6.9 (3.6; 12.1) | 6.4 (3.2; 11.4) | 5.7 (2.7; 10.5) | 4.7 (2; 9.2) |
| New Zealand | 8.5 (6.3; 11.1) | 7.6 (5.6; 10.1) | 8.2 (6.1; 10.7) | 7.4 (5.4; 9.8) | 7.7 (5.8; 10.2) | 7.8 (5.8; 10.3) | 6.3 (4.5; 8.5) | 6.4 (4.7; 8.7) | 4.9 (3.4; 6.9) | 6.3 (4.6; 8.5) |
| N. Macedonia | 9.8 (6.1; 14.8) | 10.4 (6.6; 15.7) | 6.5 (3.5; 10.9) | 5.1 (2.6; 9.2) | 7.1 (4; 11.7) | 7.6 (4.3; 12.3) | 10 (6.2; 15.3) | 7.2 (4; 11.8) | 6.7 (3.6; 11.2) | 7.6 (4.3; 12.3) |
| Norway | 2.4 (1.3; 3.9) | 1.4 (0.7; 2.7) | 1.9 (1; 3.3) | 2 (1.1; 3.5) | 2.8 (1.7; 4.4) | 2.6 (1.5; 4.2) | 3.1 (1.9; 4.8) | 2.3 (1.3; 3.8) | 2.1 (1.2; 3.6) | 2.4 (1.4; 3.9) |
| Oman | 0 (0; 0.8) | 0 (0; 0.8) | 0 (0; 0.7) | 0 (0; 0.7) | 0 (0; 0.7) | 0 (0; 0.7) | 0 (0; 0.6) | 0.3 (0; 1.2) | 0.3 (0; 1.1) | 0 (0; 0.5) |
| Singapore | 1.7 (0.8; 3.4) | 1.8 (0.8; 3.5) | 1.4 (0.5; 3) | 2.1 (1; 3.9) | 3.5 (2; 5.8) | 4.5 (2.7; 7) | 5.7 (3.6; 8.5) | 6.1 (4; 9) | 5.1 (3.1; 7.8) | 3.9 (2.2; 6.3) |
| Slovakia | 2.7 (1.5; 4.5) | 2.8 (1.5; 4.5) | 3.5 (2.1; 5.5) | 3.5 (2.1; 5.5) | 3.1 (1.8; 5) | 3.7 (2.2; 5.7) | 4.1 (2.5; 6.1) | 5.3 (3.5; 7.6) | 5.6 (3.8; 7.9) | 6 (4.2; 8.4) |
| Slovenia | 8.6 (4.9; 14) | 7 (3.7; 11.9) | 8.6 (4.9; 13.9) | 8.9 (5.2; 14.3) | 2 (0.6; 5.2) | 4 (1.7; 7.9) | 4.4 (2; 8.3) | 3.8 (1.6; 7.5) | 5.6 (2.9; 9.7) | 4.1 (1.9; 7.8) |
| South Korea | 5.1 (4.6; 5.8) | 5.9 (5.2; 6.6) | 6.6 (6; 7.4) | 7.3 (6.6; 8.1) | 7.7 (6.9; 8.5) | 7.5 (6.7; 8.3) | 7.9 (7.2; 8.8) | 7.7 (6.9; 8.5) | 7.3 (6.5; 8.1) | N/A |
| Spain | 8.6 (6.6; 11) | 9 (7; 11.4) | 9.9 (7.8; 12.4) | 9.3 (7.3; 11.7) | 10.3 (8.2; 12.7) | 8.3 (6.4; 10.5) | 9.9 (7.8; 12.2) | 9 (7.1; 11.2) | 10.5 (8.5; 12.9) | 11.6 (9.4; 14.1) |
| Sweden | 2.6 (1.7; 3.7) | 2.8 (1.9; 4) | 3 (2.1; 4.2) | 2.8 (1.9; 3.9) | 3.8 (2.8; 5.1) | 3.3 (2.4; 4.5) | 3.9 (2.9; 5.2) | 3.5 (2.6; 4.8) | 4.5 (3.4; 5.8) | 6 (4.7; 7.6) |
| UAE | N/A | N/A | N/A | 0 (0; 0.6) | 0.2 (0; 0.9) | 0.2 (0; 0.9) | 0.3 (0; 1.2) | 0.2 (0; 0.9) | 0.3 (0; 1.1) | 0.1 (0; 0.8) |
| USA | N/A | 7.4 (7.2; 7.7) | 7.1 (6.9; 7.4) | 6.9 (6.7; 7.2) | 7.1 (6.9; 7.4) | 5.8 (5.6; 6) | 5.5 (5.3; 5.7) | 4.9 (4.7; 5.1) | 5 (4.8; 5.2) | N/A |

2 SAR = special administrative region; UAE = United Arab Emirates; USA = United States of America.

**Table S5:** Clozapine utilisation prevalence (and 95% confidence intervals) per 100,000 older adults (≥65 years), 2015–2024.

| Country | 2015 | 2016 | 2017 | 2018 | 2019 | 2020 | 2021 | 2022 | 2023 | 2024 |
| --- | --- | --- | --- | --- | --- | --- | --- | --- | --- | --- |
| Austria | 196.1 (177.2; 216.5) | 196.4 (177.6; 216.7) | 197.8 (179.2; 217.9) | 194.5 (176.2; 214.2) | 180.5 (163; 199.3) | 178.7 (161.5; 197.2) | 179 (161.9; 197.3) | 182 (165; 200.3) | 170.7 (154.5; 188.2) | N/A |
| Belgium | 116 (111.3; 120.8) | 121.2 (116.5; 126.1) | 117.7 (113.1; 122.4) | 117.5 (112.9; 122.2) | 124.4 (119.7; 129.2) | 121.9 (117.3; 126.6) | 120.7 (116.2; 125.3) | 124.6 (120; 129.3) | 118.9 (114.5; 123.4) | 117.9 (113.6; 122.4) |
| Brazil | 9.3 (8.9; 9.8) | 10.1 (9.6; 10.5) | 11.1 (10.6; 11.6) | 12.5 (12; 13) | 13.6 (13.1; 14.1) | 14.1 (13.6; 14.6) | 15.7 (15.1; 16.2) | 17.8 (17.2; 18.4) | 19.8 (19.2; 20.4) | 21.8 (21.2; 22.4) |
| Brunei | 16.9 (3.5; 49.3) | 9.3 (1.1; 33.4) | 7.9 (1; 28.4) | 7.5 (0.9; 27.3) | 11.3 (2.3; 33.1) | 15 (4.1; 38.4) | 14.3 (3.9; 36.7) | 17.4 (5.7; 40.7) | 22.4 (9; 46.1) | 21.1 (8.5; 43.6) |
| Chile | 15.4 (13.7; 17.3) | 18.2 (16.3; 20.1) | 19.9 (18; 21.9) | 22 (20.1; 24.1) | 25.7 (23.6; 27.8) | 29.4 (27.3; 31.7) | 33 (30.7; 35.3) | 35.8 (33.6; 38.2) | 40.1 (37.8; 42.6) | 44 (41.5; 46.5) |
| Colombia | 397.5 (376.3; 419.5) | 360 (341.9; 378.8) | 341.8 (325.5; 358.6) | 322.8 (308; 338.1) | 280.3 (270.6; 290.3) | 291.4 (278.8; 304.3) | 269.1 (257.8; 280.8) | 250.9 (240.9; 261.2) | 241.7 (232.4; 251.3) | 215.7 (207.1; 224.5) |
| Croatia | 366.5 (353.3; 380) | 377.9 (364.6; 391.5) | 376.4 (363.2; 389.9) | 373.8 (360.8; 387.1) | 380.4 (367.4; 393.7) | 382 (369; 395.2) | 390.3 (377.3; 403.7) | 395.3 (382.3; 408.7) | 393.6 (380.7; 406.9) | 406.9 (393.9; 420.3) |
| Denmark | 36 (32.4; 39.8) | 38.3 (34.6; 42.1) | 41.3 (37.6; 45.3) | 45 (41.1; 49.1) | 43.5 (39.7; 47.5) | 42.1 (38.5; 46) | 43.6 (39.9; 47.6) | 41.5 (37.9; 45.3) | 41.1 (37.6; 44.9) | 41.3 (37.8; 45.1) |
| Ethiopia | 0 (0; 0.1) | 0.1 (0; 0.3) | 0.1 (0; 0.2) | 0.2 (0.1; 0.4) | 0.1 (0; 0.2) | 0 (0; 0.1) | 0.1 (0; 0.2) | 0.2 (0.1; 0.4) | 0.1 (0; 0.2) | 0.1 (0; 0.2) |
| Finland | 106.8 (100.9; 113.1) | 114.1 (108; 120.4) | 122.7 (116.5; 129.2) | 130 (123.6; 136.6) | 131.3 (125; 137.9) | 141.6 (135.1; 148.3) | 148.8 (142.2; 155.6) | 149 (142.4; 155.8) | 145.3 (138.9; 152) | 144.3 (137.9; 150.9) |
| France | 80.6 (79; 82.2) | 86 (84.3; 87.6) | 90.4 (88.7; 92) | 94.8 (93.2; 96.5) | 99.1 (97.4; 100.8) | 105.4 (103.7; 107.2) | 109.8 (108.1; 111.6) | 113.2 (111.4; 114.9) | 117.7 (116; 119.5) | 123.4 (121.6; 125.2) |
| Germany | 107.2 (104.6; 109.8) | 107.7 (105.1; 110.3) | 107.1 (104.5; 109.8) | 107.4 (104.8; 110.1) | 106.6 (104; 109.3) | 105 (102.4; 107.7) | 107 (104.4; 109.7) | 107.6 (105; 110.3) | 106.7 (104.1; 109.4) | 108 (105.4; 110.7) |
| Hong Kong SAR | 20 (17.5; 22.8) | 21.4 (18.8; 24.2) | 23 (20.3; 25.8) | 25.5 (22.8; 28.4) | 25.5 (22.9; 28.4) | 28.7 (25.9; 31.6) | 30.7 (27.9; 33.7) | 33 (30.2; 36) | 34.2 (31.5; 37.2) | 34.1 (31.4; 37) |
| Iceland | 327.5 (276.4; 385.4) | 323 (273; 379.4) | 292.6 (245.8; 345.7) | 289.4 (243.6; 341.3) | 301.4 (255.4; 353.3) | 272.5 (229.6; 321.2) | 348.3 (300.3; 401.8) | 357.3 (309.5; 410.5) | 328.5 (283.5; 378.7) | 339.7 (294.5; 389.7) |
| Japan | 0.4 (0.4; 0.5) | 0.5 (0.4; 0.6) | 0.5 (0.5; 0.6) | 0.6 (0.5; 0.7) | 0.7 (0.6; 0.8) | 1 (0.9; 1.1) | 1.3 (1.2; 1.4) | 1.4 (1.3; 1.6) | 1.9 (1.8; 2) | 2.4 (2.2; 2.6) |
| Latvia | 102.6 (92.7; 113.2) | 100.1 (90.4; 110.6) | 111.1 (100.9; 122.1) | 170.5 (157.8; 184) | 177.1 (164.1; 190.9) | 172.5 (159.7; 186) | 177.8 (164.9; 191.5) | 174.4 (161.6; 188) | 179.5 (166.6; 193.2) | 168.9 (156.4; 182.1) |
| Lithuania | 40.1 (35; 45.8) | 42 (36.7; 47.8) | 47.3 (41.7; 53.4) | 47.9 (42.3; 54) | 52 (46.1; 58.3) | 55.5 (49.5; 62.1) | 52.7 (46.8; 59.1) | 56.7 (50.7; 63.3) | 57 (51; 63.6) | 60.6 (54.5; 67.3) |
| Netherlands | 176 (160.9; 192.1) | 192.6 (176.7; 209.6) | 176.7 (162; 192.4) | 182 (167.3; 197.6) | 192.8 (177.8; 208.8) | 200.3 (185.2; 216.3) | 204.2 (189.4; 219.9) | 188.1 (173.9; 203.2) | 182.3 (168.4; 197) | 181.7 (168; 196.2) |
| New Zealand | 38.5 (33.9; 43.4) | 40.2 (35.7; 45.2) | 40.1 (35.7; 45) | 42.7 (38.2; 47.7) | 45.4 (40.8; 50.4) | 47.1 (42.4; 52.1) | 46.5 (42; 51.5) | 50.6 (45.9; 55.7) | 51.6 (46.9; 56.6) | 52 (47.4; 57) |
| N. Macedonia | 163.3 (148.3; 179.4) | 162 (147.3; 177.8) | 153.1 (138.9; 168.2) | 162.8 (148.5; 178.2) | 163.9 (149.7; 179.1) | 149.3 (135.9; 163.5) | 142.3 (129.4; 156.2) | 138.5 (125.9; 152) | 145.3 (132.5; 159) | 137.4 (125.2; 150.6) |
| Norway | 24.6 (21.5; 28.1) | 26.8 (23.6; 30.4) | 30 (26.6; 33.8) | 33.1 (29.6; 37) | 35.4 (31.8; 39.3) | 39.4 (35.6; 43.4) | 38.6 (34.9; 42.6) | 39.2 (35.5; 43.1) | 39.1 (35.5; 43.1) | 39.8 (36.1; 43.7) |
| Oman | 0.9 (0; 5.2) | 0.9 (0; 5) | 0 (0; 3.2) | 0 (0; 3.1) | 0 (0; 3.1) | 0.8 (0; 4.7) | 0.8 (0; 4.7) | 0 (0; 3.1) | 0 (0; 2.6) | 0 (0; 2.6) |
| Singapore | 8.3 (5.8; 11.3) | 9 (6.6; 12.1) | 11.6 (8.9; 14.9) | 12.2 (9.5; 15.5) | 13.9 (11.1; 17.3) | 13.8 (11.1; 17.1) | 14.6 (11.7; 17.8) | 17 (14; 20.4) | 18.7 (15.6; 22.1) | 19.6 (16.6; 23.1) |
| Slovakia | 77 (70.9; 83.4) | 84.1 (77.8; 90.7) | 91.4 (85; 98.1) | 97.4 (90.9; 104.2) | 101.5 (95; 108.3) | 103.3 (96.8; 110.1) | 107.9 (101.3; 114.8) | 113.9 (107.2; 120.9) | 118.5 (111.8; 125.5) | 121.7 (115; 128.7) |
| Slovenia | 172.2 (159.2; 186.1) | 170.4 (157.6; 183.9) | 162.5 (150.2; 175.6) | 165.2 (153; 178.2) | 163.7 (151.7; 176.4) | 158 (146.3; 170.3) | 139.3 (128.5; 150.8) | 138.8 (128.1; 150.1) | 135.9 (125.4; 147) | 134.3 (124; 145.3) |
| South Korea | 12.5 (11.7; 13.4) | 13.5 (12.7; 14.4) | 14.6 (13.8; 15.5) | 15.8 (14.9; 16.7) | 17 (16.1; 17.9) | 18.2 (17.3; 19.1) | 20.3 (19.4; 21.3) | 20.2 (19.3; 21.2) | 20.3 (19.4; 21.2) | N/A |
| Spain | 13.2 (11.4; 15.3) | 15.1 (13.2; 17.4) | 17 (14.9; 19.3) | 18.1 (15.9; 20.4) | 19.8 (17.6; 22.3) | 22 (19.7; 24.6) | 23.8 (21.4; 26.4) | 27.8 (25.2; 30.6) | 30.9 (28.2; 33.8) | 36 (33.1; 39.1) |
| Sweden | 66.3 (62.7; 70.1) | 68 (64.4; 71.8) | 68.2 (64.6; 72) | 69.7 (66.1; 73.5) | 70.8 (67.2; 74.6) | 72.1 (68.5; 75.9) | 71.5 (67.9; 75.2) | 70.7 (67.2; 74.4) | 69.3 (65.8; 72.9) | 71.9 (68.4; 75.5) |
| Taiwan | 45.2 (42.8; 47.7) | 48.2 (45.8; 50.7) | 54.9 (52.4; 57.5) | 56 (53.6; 58.6) | 62.8 (60.2; 65.4) | 69.6 (67; 72.3) | 71 (68.4; 73.6) | 75.1 (72.5; 77.8) | N/A | N/A |
| UAE | N/A | N/A | N/A | 0 (0; 4) | 0 (0; 3.9) | 1.1 (0; 6) | 4.1 (1.1; 10.5) | 1 (0; 5.3) | 1.8 (0.2; 6.4) | 0.8 (0; 4.6) |
| USA | N/A | 35.4 (34.9; 35.9) | 34 (33.4; 34.5) | 33.7 (33.2; 34.2) | 32.9 (32.4; 33.4) | 35.7 (35.1; 36.2) | 33.2 (32.7; 33.7) | 32.8 (32.3; 33.3) | 32.4 (31.9; 32.9) | N/A |

1 SAR = special administrative region; UAE = United Arab Emirates; USA = United States of America.

## Notes

### Author Declarations

Where applicable, the contributing authors sought ethics committee or institutional review board approval (Table S2).

